# Peripheral Epigenetic Aging Predicts Survival Beyond Brain Aging-Related Biomarkers in Centenarians

**DOI:** 10.64898/2026.05.01.26352140

**Authors:** Yaran Zhang, Marc Hulsman, Niccolò Tesi, Susan K. Rohde, Maruelle Luimes, Linda Lorenz, Sven van der Lee, Marieke Graat, Myke van der Hoorn, Dominique Daatselaar, Charlotte Teunissen, Everard G.B. Vijverberg, Alex Salazar, Henne Holstege

## Abstract

People who reach 100 have survived risks that many others have not, but which aging-related signals remain informative for their survival? We examined whether epigenetic aging provides information beyond cognition and markers of neurodegeneration in 247 Dutch centenarians recruited for reported cognitive health. Using PacBio long-read sequencing we compared five DNA methylation clocks alongside cognitive, AD-related blood biomarker, hematological measures and neuropathological assessments. GrimAge was the only clock associated with both chronological age and mortality. Higher GrimAge acceleration robustly predicted higher mortality (HR per standard deviation = 1.41, 95% CI: 1.22 - 1.63), with associations persisting after adjustment for estimated leukocyte composition and independently of MMSE. Combining GrimAge and MMSE strengthened survival stratification, with a 1.61-year difference in restricted mean survival between the lowest- and highest-risk quintiles, compared with 1.45 years for GrimAge and 1.20 years for MMSE alone. Plasma NfL was also associated with mortality but showed no detectable association with GrimAge acceleration. However, few temporally matched measurements limited assessment of their independent associations with survival. While GrimAge showed limited associations with neurodegeneration-related measures, it strongly correlated with hematological markers of aging, consistent with age-related myeloid shift. However, these did not fully account for its association with survival. Together, these findings suggest that mortality-related epigenetic signals remain informative even after exceptional survival to age 100, with blood epigenetic aging and cognition providing complementary prognostic information at extreme ages.

## Introduction

Aging involves declining physiological function and increasing susceptibility to disease. While chronological aging reflects the passage of time, biological aging reflects changes in the body’s functional state and varies among individuals of the same chronological age ^1–4^. Individuals who reach at least 100 years, centenarians, have often escaped or delayed major age-related conditions such as cardiovascular disease, cancer and dementia, which suggests that their exceptional longevity could be supported by slower biological aging ^5,6^.

Nevertheless, even among centenarians, remaining survival varies considerably. Previous work in centenarian cohorts has identified clinical markers associated with survival. In particular, those centenarians with higher scores on the Mini-Mental State Examination (MMSE), a standard measure of cognitive function, are likely to survive longer than those with lower scores^7^. In addition, elevated levels of plasma neurofilament light chain (NfL), a biomarker of prevalent neuroaxonal injury, and loss of independence in activities of daily living (ADL) predicted shorter survival in centenarians^8–11^. These measures retain prognostic value even beyond age 100.

Age related changes in DNA methylation (DNAm) have enabled the development of epigenetic clocks^12–17^. First-generation clocks were trained primarily to predict chronological age^14,18^ whereas second-generation clocks incorporated aging-related phenotypes, including clinical measures or mortality risk^19,20^. GrimAge, for example, combines DNAm-based surrogate scores for the abundance of seven plasma proteins including TIMP-1, Cystatin C, GDF-15, B2M, ADM, PAI-1, and Leptin, and a DNAm-based smoking pack-years indicator (PACKYRS) for their association with mortality. These proteins are linked to biological processes central to aging, including inflammation, metabolic adaptation, vascular aging, and immune regulation ^20^. A clock estimate that is higher or lower than expected for chronological age is described as epigenetic age acceleration or deceleration, respectively. PhenoAge- and GrimAge-based age acceleration have been associated with physical capability and the burden of chronic age-related conditions in younger and older adult populations^21–23^. Whether epigenetic clocks retain prognostic value until extreme ages, and whether clocks with different training targets remain equally informative remains unclear.

Centenarians provide an informative setting to address these questions. Centenarians have undergone exceptional survival selection, yet face annual mortality rates of approximately 40–50%^7,24^. Each additional year of survival represents substantial further selection, making differences in remaining lifespan particularly informative even within a narrow chronological-age range. Previous studies have reported DNAm age estimates substantially below chronological age in exceptionally long-lived individuals, although age-estimation performance and age-acceleration estimates differ across clocks^25–27^. These observations suggest that the relevance of the aging processes captured by clocks developed in younger populations may change with increasing age, which should be clearly observable in individuals who reached at least age 100.

Despite their shared exceptional longevity, centenarians vary considerably in DNAm age^27^, cognition and other survival-related measures. Comparing epigenetic clocks and examining their underlying signals alongside established survival markers can help determine which clocks remain informative for predicting survival, what biological processes they may reflect, and whether they provide complementary information to established markers.

To test this, we examined the blood methylation profile in 247 Dutch centenarians from the 100-plus Study, selected for reported cognitive health^28^ . Using five epigenetic clocks, the age-based clocks Hannum and Horvath, the mortality-based clocks PhenoAge and GrimAge, and the longevity-focused clock ENCen40+^14,18–20,27^, we compared the clock estimates with chronological age and assessed their associations with mortality. We then focused on GrimAge, the clock most strongly associated with mortality in this group, and examined which of its components remained associated with mortality at extreme old age. Additionally, drawing on this cohort’s extensive phenotyping, including cognitive, plasma biomarker, hematological and neuropathological assessments, we assessed whether GrimAge captures mortality-related information beyond established survival markers in centenarians.

## Results

### 1. GrimAge retains associations with chronological age and mortality in centenarians

We compared five epigenetic clocks, Hannum, Horvath, PhenoAge, GrimAge, and ENCen40+, using blood methylation profiles derived from PacBio long-read sequencing data from 247 Dutch centenarians (median age = 100 years; IQR: 100-102, Table 1), recruited into the 100-plus Study on the basis of self-reported cognitive health confirmed by a proxy^28,29^. For associations with chronological age, survival and other phenotypes, we used principal component-based versions of Hannum, Horvath, PhenoAge and GrimAge to reduce technical noise and sensitivity to missing methylation measurements^30^, while ENCen40+ was calculated using its original implementation. When analysing the component surrogates, we used the original GrimAge implementation.

**Table 1.**
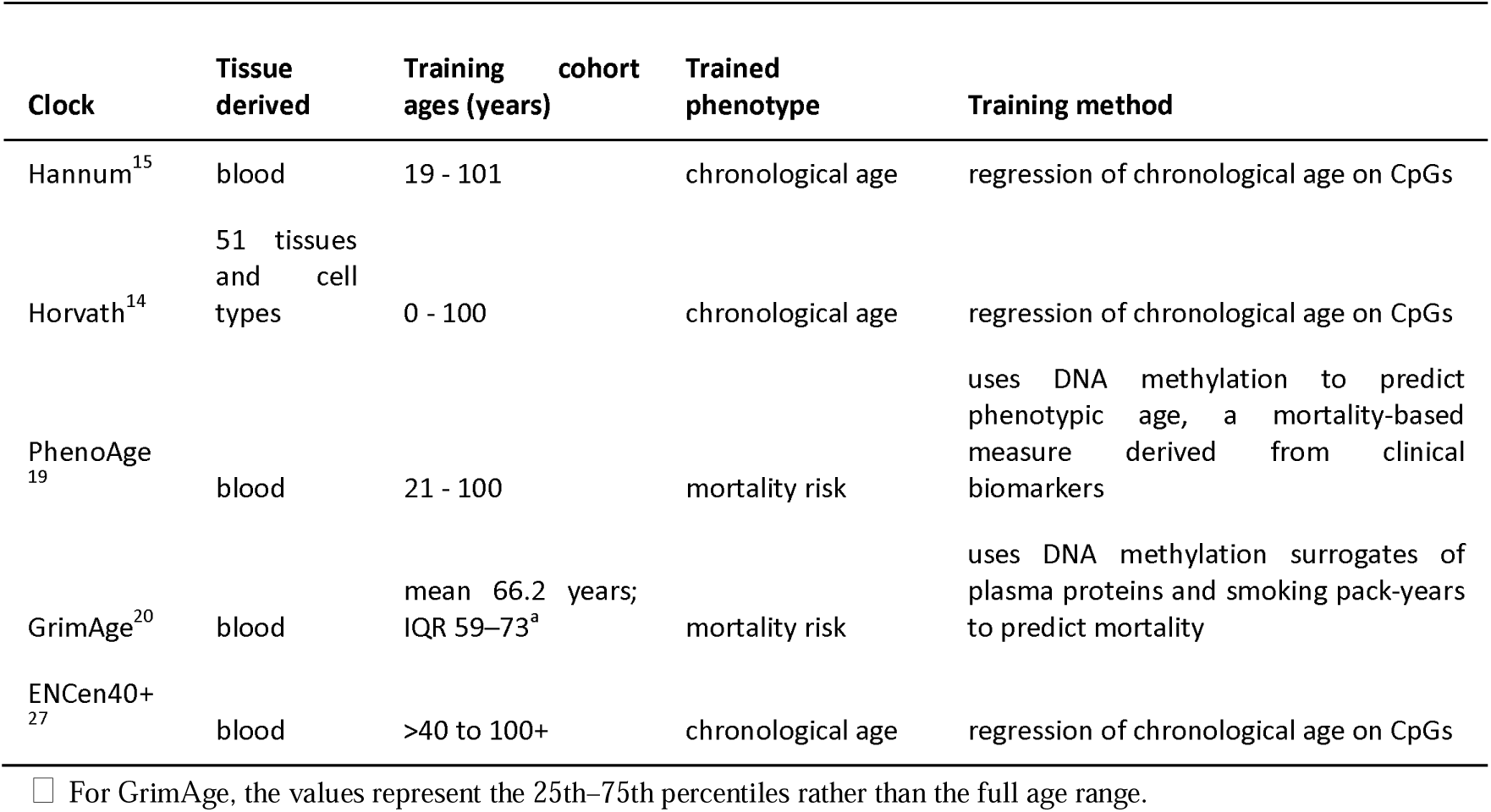
Epigenetic clocks included in this study.

Baseline blood collection of centenarians preceded death or censoring by a median of 2.1 years (IQR, 1.0–3.2). PacBio-derived methylation estimates showed high concordance with the established EPIC array measurement (Pearson r = 0.96, Supplementary Note 1, Supplementary Figure 1).

To estimate age acceleration across a broader age range, we included 246 younger Alzheimer’s Disease (AD) patients from the Amsterdam Dementia Cohort^29^ sequenced alongside the centenarians (median age = 69; IQR: 62-73). Prior studies report no consistent acceleration of blood DNAm age in AD compared with age-matched controls^31–34^ (Supplementary Note 2). All five clocks correlated strongly with chronological age in the AD cohort (Pearson r: 0.69 - 0.82) and GrimAge estimates in ten centenarian-offspring followed the same age-related trend (Supplementary Figure 6B).

Across the combined cohorts, all five clocks correlated significantly with chronological age. However, within the centenarians, only GrimAge showed a significant correlation with chronological age (Pearson r = 0.20, adj p = 1.72E-03) (Figure 1A, B, Supplementary Table 1), suggesting that GrimAge captures age-related differences within the age range of this centenarian cohort. The cross-sectional regression slope for GrimAge was shallower in centenarians than in AD patients (β = 0.48 versus 0.72), corresponding to a smaller increase in estimated GrimAge per year of chronological age (Figure 1B).

**Figure 1.**
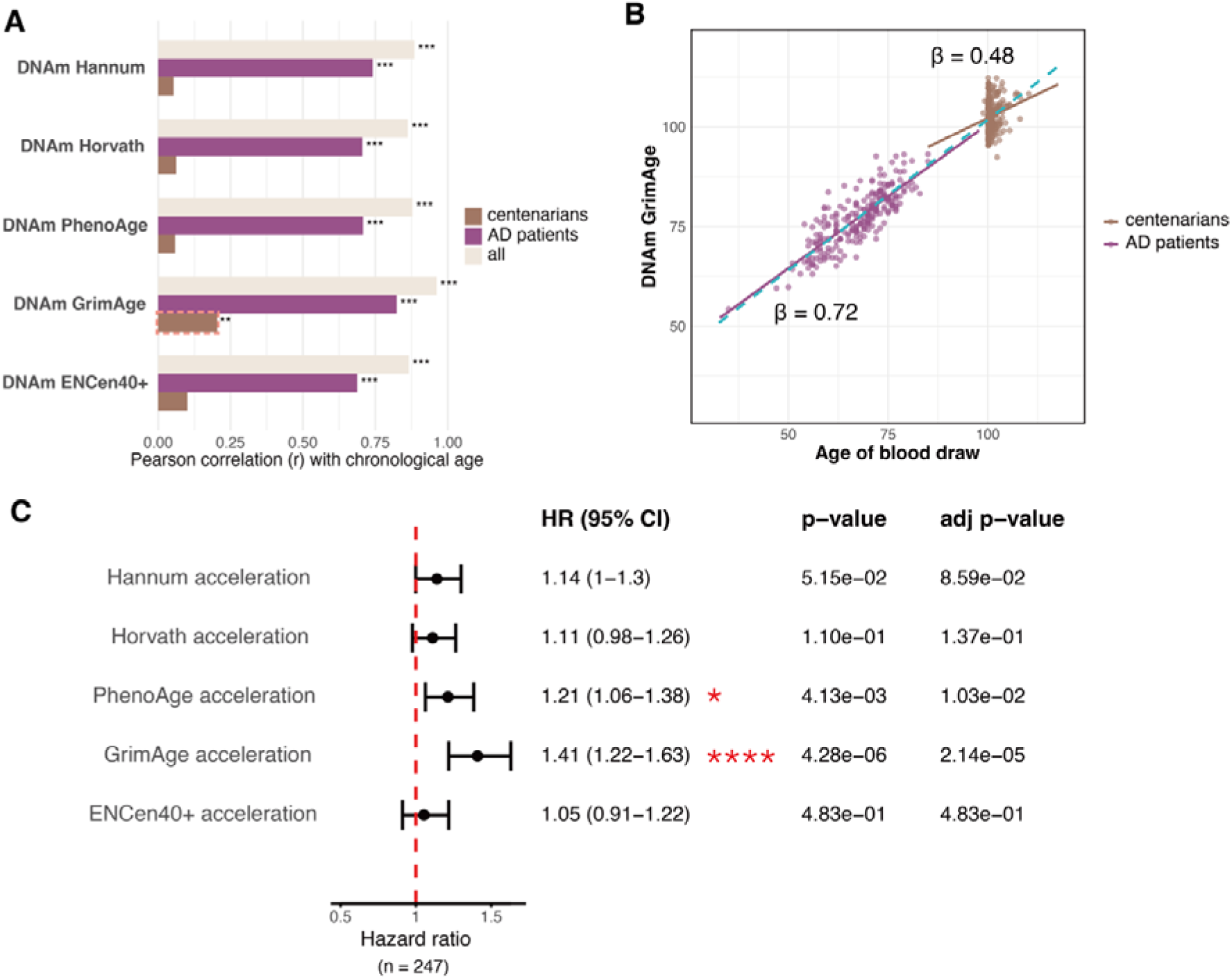
Epigenetic age and mortality associations in centenarians. **(A)** Pearson correlation between chronological age in centenarians (n= 247) and AD patients (n=246). All clocks correlate strongly with chronological age across the combined cohorts (Pearson r = 0.86-0.96; all adj. p < 5E-146). Within AD patients (Pearson r = 0.69 - 0.82; all adj. p < 5E-35). Within centenarians, only GrimAge was significantly associated with chronological age (Pearson r = 0.20, adj p = 1.72E-03). **(B)** Regression of DNAm GrimAge on chronological age across both cohorts (blue line: β = 0.76), used to calculate the age acceleration residuals. Cohort-specific regressions are shown for AD patients (purple line: β = 0.72), and centenarians (brown line: β = 0.48). **(C)** Mortality associations of age accelerations derived from five clocks in 247 centenarians (228 deaths, 19 censored). Cox models were adjusted for sex and 5 genetic PCs. Hazard ratios represent a one-standard deviation increase in age acceleration.

Across cohorts, GrimAge acceleration did not differ significantly between centenarians (median = −0.12 years; IQR: −2.44 to 2.53) and AD patients (median = −0.68 years; IQR: −2.81 to 2.40; Wilcoxon p = 4.06E-01). We additionally assessed whether centenarians’ GrimAge estimates deviated from the extrapolated age trend in AD patients. Estimates were on average 1.25 years above this trend, but the deviation was not statistically significant (95% bootstrap CI: −0.77 to 3.28; Figure 1B).

We calculated age acceleration as the residual from regressing each clock’s DNAm age estimate on chronological age across both cohorts (Figure 1B). In Cox models adjusted for sex and genetic principal components, PhenoAge- and GrimAge-acceleration were significantly associated with mortality (PhenoAge acceleration: HR = 1.21, 95% CI: 1.06 - 1.38, adj p = 1.03E-02; GrimAge acceleration: HR = 1.41, 95% CI: 1.22 - 1.63, adj p = 2.14E-05) (Figure 1C), consistent with the mortality-oriented design of these clocks. These estimates represent respectively a 21% and 41% higher mortality hazards per standard deviation increase. Centenarian-only models including unresidualized DNAm age and chronological age as separate terms yielded similar associations (GrimAge: HR = 1.40, 95% CI: 1.21–1.63, adjusted p = 3.55E-05; PhenoAge: HR = 1.20, 95% CI: 1.05–1.37, adjusted p = 1.76E-02; Supplementary Figure 6A), indicating that these findings did not depend materially on the AD calibration cohort.

While additional adjustment for methylation-estimated leukocyte fractions attenuated GrimAge mortality association, it nevertheless remained significant (HR = 1.28, 95% CI: 1.08–1.53, p = 4.70E−03, adjusted p = 2.35E−02), whereas none of the other clocks remained significant after multiple-testing correction (Supplementary Figure 7A). Several estimated leukocyte fractions were themselves associated with mortality, with lower CD4+ T-cell and B-cell fractions and a higher neutrophil fraction associated with higher mortality (Supplementary Figure 7C), consistent with blood-cell composition contributing to the mortality-related signal captured by GrimAge^20,35,36^. We therefore focused subsequent analyses on GrimAge, the only clock associated with both chronological age and mortality and retaining a mortality association after leukocyte adjustment within the centenarian cohort.

Because GrimAge is a composite measure derived from DNAm-based surrogates for plasma protein abundance and smoking pack-years, we also examined the relationships among its component scores in centenarians. Most DNAm components were intercorrelated, and GrimAge correlated with most component scores, although correlations with DNAmPAI1 and DNAmGDF15 were weaker (Supplementary Figure 5, Supplementary Data 2). DNAmTIMP1, DNAmCystatinC, DNAmB2M and DNAmPACKYRS were each associated with mortality after multiple testing correction (Supplementary Figure 2A). Leukocyte adjustment attenuated these associations, only DNAmTIMP1 remained significant after correction (Supplementary Figure 2B). Component-level associations with cognitive, neuropathological, plasma biomarker and other phenotypes were generally limited (Supplementary Note 3, Supplementary Data 1).

### 2. GrimAge predicts mortality independently of MMSE and shows no association with plasma NfL

We compared GrimAge acceleration with established survival markers Mini-Mental State Examination (MMSE) scores, Barthel Index (BI) and plasma neurofilament light chain (NfL) in 132 centenarians with all four measures available (121 deaths, 11 censored; Table 2, Figure 2A). MMSE and BI were assessed on the day of the blood draw used for GrimAge estimation, a median of 2.5 years before death or censoring (IQR: 1.8-3.7 years). Plasma NfL was measured a median of 1.03 years later (IQR: 0.02-1.83 years), and a median 1.34 years before death or censoring (IQR 0.68-2.55 years).

**Figure 2.**
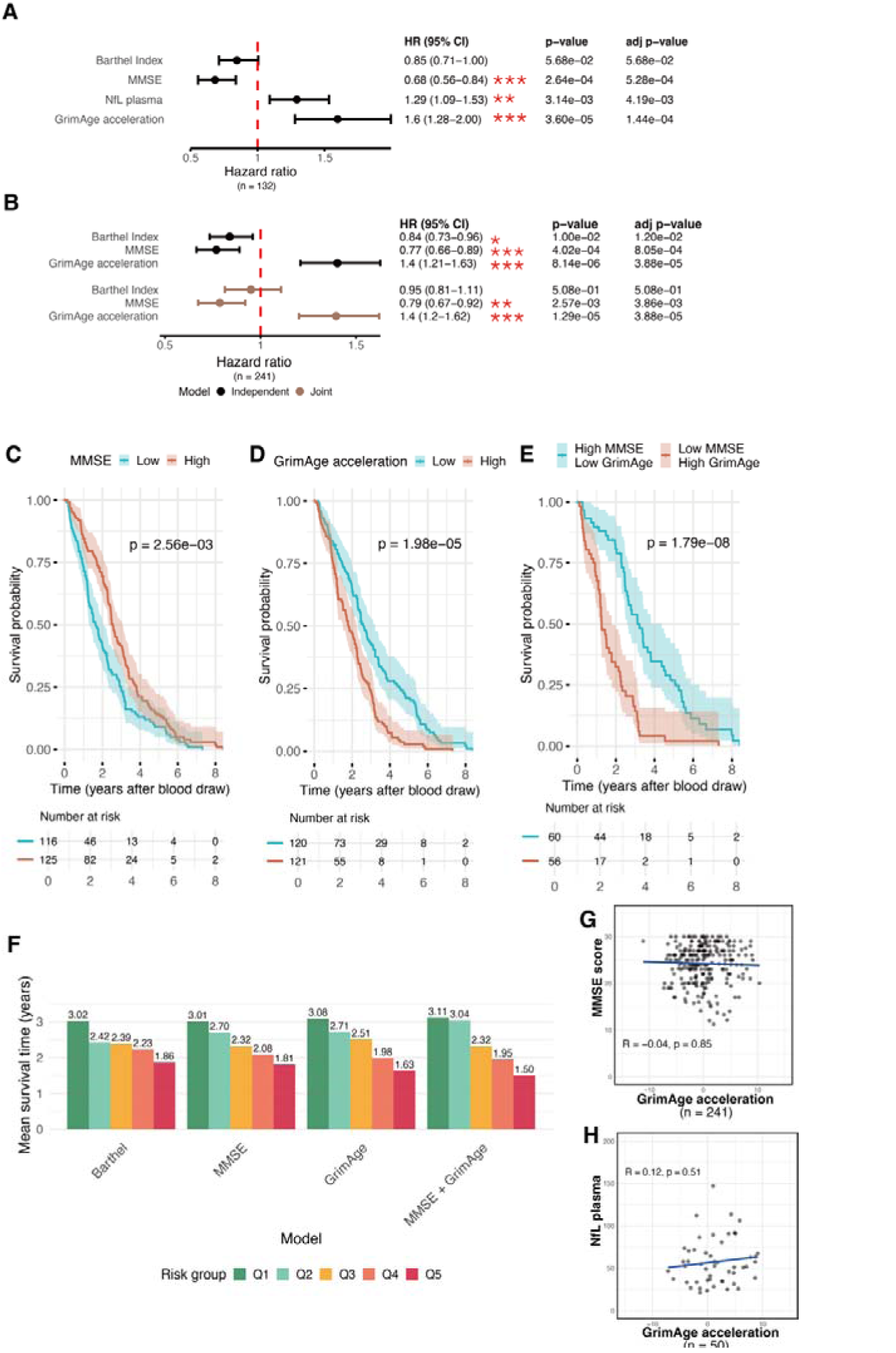
Survival associations of GrimAge acceleration, MMSE score, BI and plasma NfL. **(A)** Cox regression models in 132 centenarians with all four measures available. Models were adjusted for age, sex, education attainment and genetic PCs. All variables were z-score standardized before analysis, such that hazard ratios represent the effect of a 1 SD increase in each variable. MMSE and Barthel Index were assessed on the same day as the collection of the blood sample used for GrimAge estimation; NfL sampling occurred a median of 1.03 years later. Survival was measured from the respective assessment or sampling date. (B) Separate (black) and joint (brown) Cox models for concurrently measured MMSE, BI and GrimAge acceleration (n = 241). Models were adjusted for age, sex, education and 5 genetic principal components. (C-E) Kaplan-Meier survival curves were generated based on the median split of (C) MMSE score of 25, (D) GrimAge acceleration of -0.09, and (E) MMSE combined with GrimAge acceleration in centenarians. (F) RMST over an 8.4-year follow-up horizon across quintiles of Cox-predicted mortality risk from models including BI, MMSE, GrimAge acceleration, or MMSE and GrimAge acceleration together. Values above each bar indicate RMST in years. (G) Association between GrimAge acceleration and imputed MMSE scores (n = 241), with the education-adjusted partial Pearson correlation and FDR-adjusted p-value. (H) Association between GrimAge acceleration and plasma NfL in individuals sampled within <0.2 years for both measures (n = 50), with the Pearson correlation and FDR-adjusted p-value. Blue lines in G-H show fitted linear regressions.

**Table 2.** Baseline characteristics of centenarian subsets.

|  | MMSE + BI + NfL + GrimAge subset<br>(n = 132) | MMSE + BI + GrimAge subset<br>(n = 241) |
| --- | --- | --- |
| Deaths, n (%) | 121 (91.7%) | 223 (92.5%) |
| Censored, n (%) | 11 (8.3%) | 18 (7.5%) |
| Age at baseline, years, mean ± SD | 100.86 ± 1.15 | 101.13 ± 1.45 |
| Female, n (%) | 88 (66.7%) | 168 (69.7%) |
| MMSE score, mean ± SD | 25.21 ± 3.56 | 24.25 ± 4.11 |
| BI (self-reported), mean ± SD | 16.22 ± 3.35 | 15.71 ± 3.38 |
| plasma NfL (pg/ml), median [IQR] | 52.70 [37.57–74.62] | — |

Values are presented as mean ± SD unless otherwise indicated. Plasma NfL concentrations are presented as median [IQR]. In the NfL subset (n = 132), plasma NfL was measured a median of 1.03 years [0.02–1.83] after MMSE/BI/GrimAge assessment.

In independent Cox regression models adjusted for age, sex, education and genetic PCs, MMSE (HR = 0.68, 95% CI: 0.56 - 0.84, adj p = 5.28E-04) and plasma NfL (HR = 1.29, 95% CI: 1.09–1.53, adj p = 4.19E-03) were significantly associated with mortality. BI did not show significant association with mortality in this subset of centenarians (HR = 0.85, CI: 0.71-1.00, adj p = 5.68E-02). Notably, GrimAge acceleration showed a stronger association with mortality than both MMSE and plasma NfL with a HR of 1.60 (95% CI: 1.28–2.00, adj p = 1.44E-04) in this subset.

We further examined the independent associations of concurrently measured MMSE, BI and GrimAge acceleration in 241 centenarians (223 deaths, 18 censored; Table 2, Figure 2B). In a joint Cox model, MMSE (HR = 0.79, 95 % CI 0.67 - 0.92, adj p = 3.86E-03) and GrimAge acceleration (HR = 1.40, 95 % CI 1.20 - 1.62, adj p = 3.88E-05) remained significantly associated with mortality, whereas the BI did not (HR = 0.95, 95 % CI 0.81 - 1.11, adj p = 5.08E-01). This suggests that MMSE and GrimAge acceleration are both independent and significant components contributing to mortality risk, whereas the BI does not provide additional predictive value when these factors are considered.

Next, we stratified the 241 centenarians by MMSE (≥25 versus <25; cohort median = 25) and GrimAge acceleration (above vs. below the cohort median, -0.09) and compared survival across groups. In line with our previous results^7^, individuals with MMSE ≥25 showed lower mortality (log-rank p = 2.56E−03) (Figure 2C), whereas individuals with GrimAge acceleration above the cohort median had higher mortality (log-rank p = 1.98E−05) (Figure 2D). Additionally, for both MMSE and age acceleration, we calculated the restricted mean survival time (RMST) over the longest observed follow-up (8.4 years). In the lowest 20% Cox-predicted risk stratum, the RMST for MMSE was 3.01 years, compared with 1.81 years in the 20% highest-risk stratum. For GrimAge acceleration, the corresponding RMSTs were 3.08 and 1.63 years, respectively (Figure 2F). These results indicate that both cognitive status and epigenetic age acceleration effectively, and with similar effect sizes, stratify survival at extremely old ages.

A stratified survival analysis further indicated that individuals with low MMSE scores combined with higher GrimAge acceleration (low-MMSE and high-GrimAge, n=56), had significantly worse survival outcomes compared to the high-MMSE and low-GrimAge group (n = 60) (log-rank test, p = 1.79E-08) (Figure 2E, Supplementary Figure 3C). To quantify the joint prognostic effect of both markers, we fitted a Cox model that included MMSE and GrimAge acceleration together while adjusting for chronological age, sex, education, and the genetic PCs. Over the full 8.4-year horizon, the RMST was 3.11 years in the lowest 20% risk stratum and 1.50 years in the highest 20% risk stratum (Figure 2F). Combining MMSE and GrimAge acceleration distinguished low- and high-risk groups more clearly than either marker alone, supporting their complementary value for survival stratification (Figure 2F). Consistent with their independent contributions to mortality risk, MMSE scores showed no association with GrimAge acceleration (partial Pearson r = –0.04, adj p = 0.85) (Figure 2G).

In time-stratified Cox regression analyses, both MMSE and GrimAge acceleration were significantly associated with mortality within the first year (n = 239; MMSE: HR = 0.93, 95% CI: 0.87–1.00, adj p = 4.72E−02; GrimAge acceleration: HR = 1.09, 95% CI: 1.01–1.18, adj p = 4.31E−02) and during years 1–3 (n = 189; MMSE: HR = 0.93, 95% CI: 0.89–0.97, adj p = 6.72E−03; GrimAge acceleration: HR = 1.08, 95% CI: 1.02–1.15, adj p = 1.12E−02). Beyond 3 years (n = 71), GrimAge acceleration showed its largest hazard ratio and most significant association (HR = 1.16, 95% CI: 1.07–1.26, adj p = 1.34E−03), while MMSE was no longer significantly associated with mortality (HR = 1.01, 95% CI: 0.93–1.09, adj p = 8.39E−01) (Supplementary Figure 3D). This suggests that MMSE more strongly associates with shorter-term survival, while GrimAge acceleration has a stronger association with longer-term survival dynamics.

For plasma NfL, survival did not differ significantly between groups defined by a median split (52.70 pg/mL) in the subset with NfL, GrimAge, MMSE and BI data available (n = 132; log-rank test, p = 8.14E-01). However, in the larger set of centenarians with available plasma NfL (n = 252), a tertile-based analysis revealed that individuals in the highest plasma NfL tertile (64.30 - 378.65 pg/mL) had significantly shorter survival (log-rank test, p = 2.97E-03), with mean survival times of 1.4 and 2.0 years in the highest and lowest tertiles, respectively (Supplementary Figure 3A-B). Only 50 individuals had NfL and GrimAge measurements within 0.2 years, limiting precision in their joint survival model (Supplementary Figure 3E). However, in this temporally matched subset, NfL showed no detectable correlation with GrimAge acceleration (Pearson r = 0.12, adjusted p = 0.51; Figure 2H).

### 3. GrimAge acceleration associates with hematological markers, but these do not fully explain its association with survival

Because GrimAge predicted survival independently of established markers such as MMSE, we examined biological processes that might underlie its signal. GrimAge combines DNAm-based surrogates of plasma proteins and smoking exposure selected for their associations with mortality^20^. We therefore correlated GrimAge acceleration with hematological markers in 133 centenarians with both DNAm and blood measurements (Figure 3A, Supplementary Data 2). Markers in Figure 3A are ordered based on hierarchical clustering of their intercorrelations, such that markers reflecting shared biological processes appear together (Supplementary Figure 4A, Supplementary Data 2).

**Figure 3.**
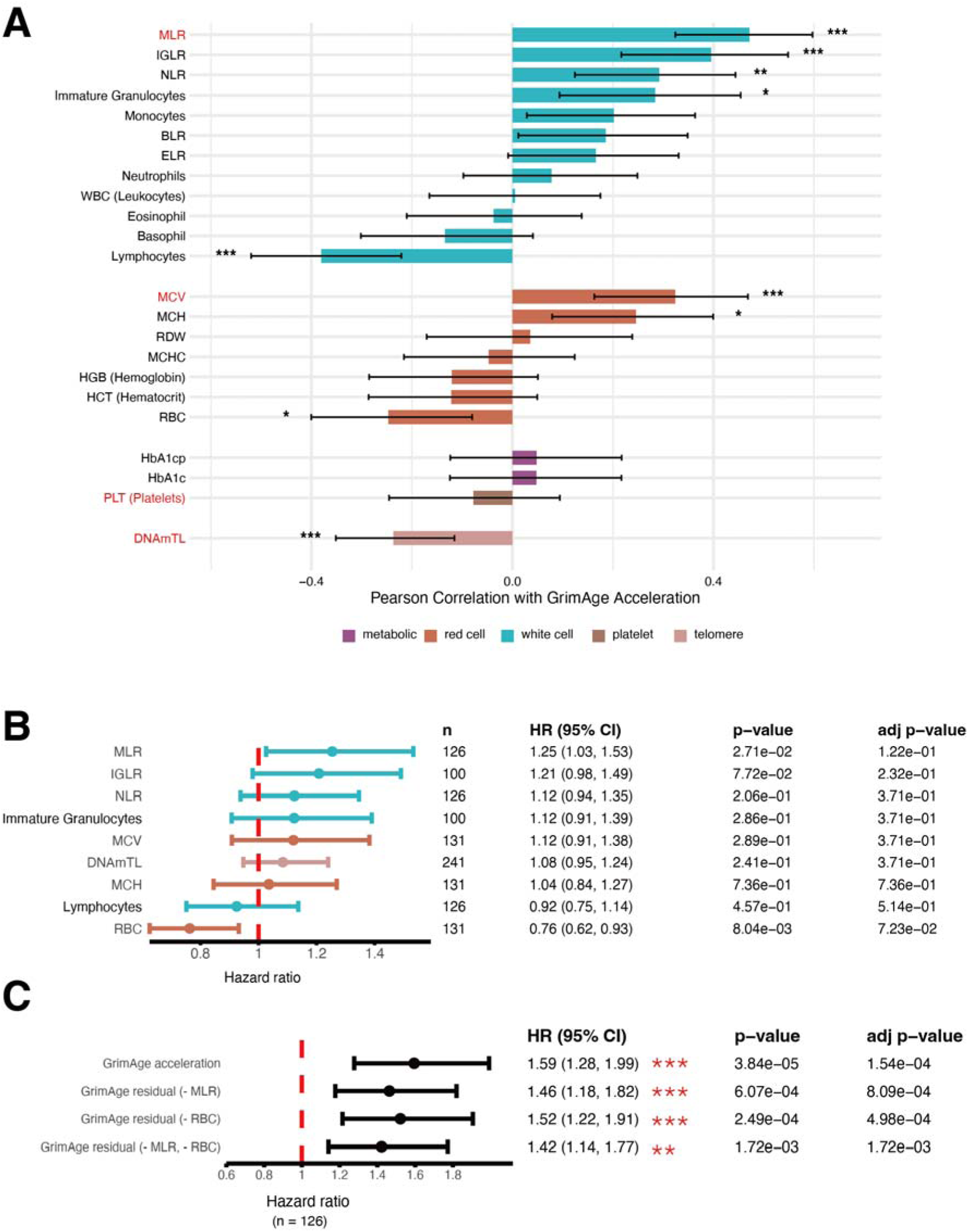
Associations of GrimAge acceleration with hematological markers and mortality. (A) Pearson correlations with hematological markers (n = 133) and DNAm-estimated telomere length (DNAmTL; n = 247). Error bars indicate 95% confidence intervals. Markers are ordered by hierarchical clustering of their intercorrelations (Supplementary Figure 4A) and labeled by class: metabolic, red cell, white cell, platelet and telomere. (B) Cox regression survival analysis for hematological markers significantly associated with GrimAge acceleration. HRs represent the effect of a 1 SD increase in each variable. (C) Cox regression survival analysis examining associations between survival and GrimAge acceleration, GrimAge residuals after regressing out MLR (GrimAge residual (− MLR)), after regressing out RBC (GrimAge residual (− RBC)), and after regressing out both MLR and RBC (GrimAge residual (− MLR, − RBC)) (n = 126). All models were adjusted for age, sex and 5 genetic PCs.

Across the hematological panel, GrimAge acceleration correlated most strongly with markers consistent with the age-associated “myeloid shift,” an increasing proportion of myeloid cells in peripheral blood^37,38^. It showed a negative association with lymphocyte count (Pearson r = −0.38, adj p = 1.12E−04), together with strong positive associations with myeloid/lymphoid ratios: monocyte-to-lymphocyte (MLR; Pearson r = 0.47, adj p = 4.75E−07), immature granulocyte-to-lymphocyte (IGLR; Pearson r = 0.40, adj p = 3.13E−04), and neutrophil-to-lymphocyte (NLR; Pearson r = 0.29, adj p = 3.77E−03). GrimAge acceleration also correlated positively with immature granulocyte count (Pearson r = 0.28, adj p = 1.19E−02), mean corpuscular volume (MCV; Pearson r = 0.32, adj p = 7.77E−04), and mean corpuscular hemoglobin (MCH; Pearson r = 0.25, adj p = 1.19E−02), and negatively with red blood cell count (RBC; Pearson r = −0.25, adj p = 1.19E−02). DNAmTL, a methylation-based estimator of leukocyte telomere length, correlated modestly and negatively with GrimAge acceleration (Pearson r = −0.24, adj p = 7.9E−04), suggesting partially distinct biological signals^39^.

Using the hierarchical clustering structure (Supplementary Figure 4A, Supplementary Data 2), we selected MLR, MCV, platelet count (PLT), and DNAmTL as representatives of the major clusters because they showed the strongest associations with GrimAge acceleration within their respective clusters (see Methods). MLR, MCV, and DNAmTL remained significant in a joint regression model, suggesting partially independent contributions to GrimAge acceleration (Supplementary Figure 4B, Supplementary Data 2).

We next assessed whether these GrimAge-associated hematological markers were associated with survival in the 133 centenarians with hematological markers available. Despite their correlations with GrimAge acceleration, none of the inflammatory or myeloid-associated markers remained significant after multiple-testing correction (Figure 3B), while several methylation-estimated leukocyte fractions were associated with survival in the full cohort (n = 247; Supplementary Figure 7C). Only MLR and RBC showed nominal associations with survival (MLR: HR = 1.25, 95% CI: 1.03–1.53; RBC: HR = 0.76, 95% CI: 0.62–0.93).

Given MLR’s established links to immune aging and chronic inflammation^37,38^ and the nominal survival associations of MLR and RBC, we tested to what extent these markers accounted for GrimAge’s survival association. In a Cox model including GrimAge acceleration and these markers (n = 126; 116 deaths), GrimAge acceleration remained associated with reduced survival (HR = 1.48, 95% CI: 1.15–1.91, p = 2.50E−03), whereas MLR (HR = 1.10, 95% CI: 0.86–1.40, p = 4.39E−01) and RBC (HR = 0.81, 95% CI: 0.65– 1.01, p = 6.28E−02) did not (Supplementary Figure 4C). Consistently, GrimAge residuals also remained associated with reduced survival after regressing out MLR (HR = 1.46, 95% CI: 1.18–1.82, adj p = 8.09E−04), RBC (HR = 1.52, 95% CI: 1.22–1.91, adj p = 4.98E−04), or both (HR = 1.42, 95% CI: 1.14–1.77, adj p = 1.72E−03; Figure 3C). Thus, GrimAge acceleration associated with survival beyond these hematological markers.

Although DNAmTL correlated modestly and inversely with GrimAge acceleration, it was not associated with survival (HR = 1.08, 95% CI: 0.95–1.24, adj p = 3.71E−01; Figure 3B). This is consistent with telomere-related variation contributing only modestly to GrimAge acceleration and not accounting for its association with mortality risk in this centenarian cohort^20^.

### 4. GrimAge acceleration shows limited association with brain aging and other age-related phenotypes

To identify phenotypes that might underlie the association between GrimAge acceleration and mortality, we tested GrimAge against a broad panel of neuropathological measures (available for n = 62 participants), cognitive performance (n = 241), AD related plasma biomarkers (n = 50), BMI (n = 188), education (n = 241), smoking history (n = 238) and other medical history records (n = 241) (Supplementary Note 3; Supplementary Tables 2-6). After correction for multiple testing, only lower cortical Aβ40 load in Brodmann area 38 (BA38) remained significantly associated with higher GrimAge acceleration, whereas most other phenotypes, including cardiovascular conditions, showed no association, and the few nominal associations did not survive multiple-testing correction and had limited effect sizes. GrimAge acceleration therefore showed limited detectable associations with these phenotypes in this cohort.

## Discussion

Among five epigenetic clocks, GrimAge was the only clock associated with both chronological age and mortality in centenarians, and the only clock retaining a significant mortality association after leukocyte adjustment. GrimAge also remained associated with mortality independently of MMSE, demonstrating that it captures mortality-related information beyond cognitive status even after age 100.

Previous work reported epigenetic age estimates below chronological age in exceptionally long-lived individuals including semi-supercentenarians^40^. Younger patients with AD and centenarians selected for reported cognitive health represent strongly contrasting cognitive outcomes for their ages. Yet centenarians did not show downward shift in GrimAge relative to the extrapolated trend in younger AD patient. Exceptional past survival therefore need not translate into lower current values of a mortality-trained clock. We did observe a shallower cross-sectional slope within centenarians when regressing GrimAge to chronological age within centenarians compared to AD patients. This could reflect slower accumulation of methylation changes after reaching extreme old age, selective survival of individuals with lower GrimAge, or differences in clock calibration at these ages. Further studies using cognitively healthy reference cohorts and longitudinal methylation data are needed to confirm these patterns and clarify how they develop as individuals approach and survive beyond age 100.

GrimAge remained associated with mortality independently of MMSE, showing that it provides survival information independent of cognitive status. In addition, GrimAge also shows limited detectable overlap with the available neurodegeneration-related measures in this cohort. Mortality was highest among centenarians with both high GrimAge acceleration and low MMSE scores, and combining them improved survival stratification. Although both markers were associated with mortality during earlier follow-up, MMSE showed its strongest evidence of association more proximal to sample collection, whereas GrimAge acceleration showed its strongest association over longer follow-up. This pattern suggests that cognition may reflect more immediate vulnerability, while GrimAge acceleration also captures processes relevant to longer-term survival. In contrast, the BI did not retain independent predictive value once MMSE and GrimAge were included, suggesting overlapping prognostic information. One possible explanation is that physical independence partly reflects downstream consequences of cognitive impairment and systemic aging, rather than an entirely separate mortality domain in this population.

To explore which GrimAge-related signals remain informative after age 100, we examined the original clock’s DNAm protein and smoking surrogates. Compared with the original GrimAge study in younger individuals, fewer DNAm components were significantly associated with mortality in centenarians^20^. The retained associations of DNAmTIMP1, DNAmB2M and DNAmCystatinC suggest links to tissue remodeling and immune–renal physiology^41–43^. Of these, only DNAmTIMP1 remained significant after leukocyte adjustment and multiple-testing correction. For other protein-surrogate components, related to stress responses, vascular regulation and metabolism, we did not observe significant associations with mortality^44–47^. This suggests that the physiological vulnerabilities associated with mortality may shift with aging until extreme old age, potentially reflecting selection for resilience to vascular and metabolic dysfunction among individuals reaching age 100^28,48^. DNAmPACKYRS, the component most significantly associated with mortality in the original study^20^, was also associated with mortality in our cohort, although this association did not survive multiple-testing correction after leukocyte adjustment. Moreover, DNAmPACKYRS showed no detectable association with reported smoking exposure. Its mortality association among never-smokers in the original study suggests that it captures information beyond reported smoking^20^. Survival selection may further influence this relationship: individuals who reach 100 despite smoking may be enriched for protective factors that reduce their susceptibility to smoking-related harm. These analyses assess which individual GrimAge surrogates remain associated with mortality at extreme ages, rather than quantify their contributions to the overall clock’s mortality association. These surrogate analyses provide biological context for GrimAge’s association with survival, although they may not fully reflect the signals captured by the principal component-based version used in our main analyses.

Unlike findings in some broader aging cohorts^21,49–51^, GrimAge acceleration showed no detectable association with cognitive performance. Plasma biomarker and neuropathological associations were also limited, apart from an inverse association with cortical Aβ40. Despite recruitment for self-reported cognitive health confirmed by a proxy^52^, participants varied substantially in cognition at the time of testing and subsequent survival. Smoking history, education and BMI likewise showed no significant associations with GrimAge acceleration, despite reported associations in younger populations^53,54^. Survival to age 100 may enrich for protective factors that offset risks associated with lifestyle and socioeconomic circumstances, potentially attenuating associations observed in broader aging cohorts.

GrimAge acceleration showed a clearer overlap with hematological aging, correlating with lower lymphocyte counts and higher myeloid-to-lymphocyte ratios, consistent with an age-related myeloid shift^37,38^. This shift may partly reflect age-related clonal hematopoiesis (ARCH), which can be extensive in centenarians, with expanded clones contributing disproportionately to myeloid cells^55,56^. We previously documented pronounced clonal expansion in women aged 111 and 115 years, including preferential contribution to myeloid cells in the former^55,56^. Common ARCH driver-genes DNMT3A and TET2 regulate DNA methylation, and mutations in these genes have been associated with altered methylation at clock CpGs and epigenetic age acceleration^57^. Expansion of clones with altered methylation profiles could therefore also contribute to the blood-based aging signals captured by GrimAge. Leukocyte adjustment reduced the mortality hazard ratio for GrimAge acceleration from 1.41 to 1.28 per standard deviation, although the association remained significant. The degree of attenuation could reflect a greater contribution of hematological aging at extreme old age, although this requires confirmation through direct comparisons with younger cohorts. The association also persisted after adjustment for measured MLR and RBC. Together, these findings suggest that GrimAge at least in part overlaps with immune and hematological aging, while also capturing broader systemic aspects of aging relevant to survival at extreme old age.

High concordance between PacBio-derived and EPIC-array methylation measurements supports applying established clocks to long-read sequencing data. Long reads additionally provide sequence variants, structural variants and haplotypes, creating opportunities to connect inherited variation, clonality and allele-specific methylation with epigenetic aging^58–60^.

A distinctive strength of our centenarian cohort is the depth of phenotyping in individuals who have survived exceptionally long yet face high subsequent mortality. Concurrent cognitive assessment and blood methylation profiling, linked to survival follow-up, hematological and plasma measurements, and *post-mortem* neuropathology in a subset, allowed us to examine epigenetic aging signals alongside these measures. While such linked data are rare in centenarian research, modest sample sizes limit statistical power to detect associations. For example, while plasma NfL was associated with mortality in 252 centenarians, only 50 had temporally matched plasma NfL and GrimAge measurements, limiting assessment of whether they are independent markers of mortality risk. The similar mortality associations obtained with and without AD-based calibration suggest that this result was not driven by the choice of reference cohort. However, the single-cohort design and relatively homogeneous population warrant replication in centenarian cohorts with different population backgrounds and recruitment criteria. Independent cohorts would also help assess the robustness of slope estimates, given the substantial interindividual variation and fewer observations at the oldest ages. Intercorrelations among leukocyte fractions and among GrimAge components make their independent contributions difficult to disentangle. Although these observational analyses cannot establish causal mechanisms, they generate testable biological hypotheses about the processes that remain relevant to survival after age 100.

In conclusion, GrimAge is a robust predictor of mortality in centenarians recruited for reported cognitive health and retains prognostic value independently of MMSE, with their combination strengthening survival stratification. GrimAge overlapped with hematological aging processes while showing limited associations with most brain aging-related measures, including plasma NfL. These findings indicate that peripheral epigenetic aging captures mortality-related biological changes beyond cognitive status that remain relevant even at extreme old age. The GrimAge surrogates associations suggest that the importance of mortality-related epigenetic signals shift with advancing age and increasing survival selection.

## Methods

### Study cohort

For DNA methylation profiling we used peripheral blood samples collected from 247 centenarians from the 100-plus Study recruited for reported cognitive health^28^. Detailed information on the 100-plus Study, including cohort design, inclusion and exclusion criteria, baseline and follow-up assessments, general characteristics, and mortality data, is available in Holstege et al.^28^. Of these, 228 (92.3%) died and 19 (7.7%) were censored. Centenarians selected for long-read sequencing were prioritized based on health and sample availability criteria, including European ancestry, unrelatedness, baseline MMSE >25, PBMC availability, and availability of familial DNA and brain donation. As a calibration cohort we used the genomes derived from peripheral blood samples from 246 Alzheimer’s Disease (AD) patients, that were sequenced simultaneously with the centenarians. AD patients were selected from the Amsterdam Dementia Cohort^29^. AD samples were selected based on similar criteria, including European ancestry, unrelatedness, PBMC availability, sex balance, and brain availability, with final inclusion constrained by DNA availability. Ten offspring of centenarians, whose peripheral-blood samples were processed using the same PacBio long-read sequencing pipeline, were also included.

The 100-plus Study and the Amsterdam Dementia Cohort were conducted in accordance with the ethical principles of the Declaration of Helsinki. The 100-plus Study was approved by the Medical Ethics Review Committee of VU University Medical Center, now Amsterdam UMC (METC registration number 2016.440; national study number NL57750.029.17), and the 100-plus Study Biobank was approved under registration number 2017.122. The Amsterdam Dementia Cohort, including the storage and sequencing of DNA, was approved by the Medical Ethics Review Committee of VU University Medical Center (approval numbers 2016.061 and 2017.315). All participants provided informed consent for participation and the use of their data and biological materials for scientific research.

Post-mortem brain donation procedures were carried out using services provided by the Netherlands Brain Bank under protocol 2009/148; the resulting brain tissues form part of the 100-plus Study brain collection. Further information on the ethical approval and consent procedures for the 100-plus Study is available at https://holstegelab.eu/ethics/.

### DNA methylation and epigenetic clocks

For centenarians, blood samples were collected during baseline study visits. DNA methylation was profiled using PacBio HiFi long-read sequencing technology. Details of the sequencing procedures and data processing are described in Salazar et al.^61^. Methylation scores were generated using PacBio’s pb-CpG-tools (version 2.3.2, https://github.com/PacificBiosciences/pb-CpG-tools) with default parameters.

To validate PacBio-derived methylation estimates, one centenarian sample with a global sequencing coverage of 46.87× was also analyzed using the Illumina Infinium MethylationEPIC BeadChip array. Raw EPIC files were processed using minfi (version 1.48.0), with Noob background correction and dye-bias normalization (preprocessNoob). CpG coordinates were intersected between platforms, and concordance of methylation β-values at shared sites was assessed using Pearson correlation. To examine the effect of read depth, we randomly selected 10,000 shared CpG sites and subsampled 5, 10, 15, and 20 reads per site. Methylation estimates were recalculated using pb-CpG-tools and compared with the corresponding EPIC β-values using Pearson correlation. Validation results are presented in Supplementary Note 1 and Supplementary Figure 1.

Epigenetic clock estimates were derived from the DNA methylation data from PacBio sequencing. To further reduce technical noise and the impact of missing values, we employed principal component–based versions of the Hannum, Horvath, PhenoAge, and GrimAge clocks, which are more robust to noise and missing data, using the dnaMethyAge package (https://github.com/yiluyucheng/dnaMethyAge), with PCHannum, PCHorvath, PCPhenoAge, and PCGrimAge. DNAm ENCen40+ was estimated using the Centenarian-Clock tool (https://github.com/victorychain/Centenarian-Clock).

### DNAm components

DNA methylation components of GrimAge, representing estimated plasma protein levels, were computed using the meffonym package (https://github.com/perishky/meffonym). For correlation analyses, DNAm components were adjusted for chronological age by regressing out age effects prior to testing associations.

To examine the internal structure of DNAm GrimAge and its component methylation-based surrogates, we computed pairwise Pearson correlations among DNAm GrimAge, its 8 DNAm components, chronological age, and sex in the centenarian cohort. Correlation significance was assessed using two-sided tests with FDR correction. Hierarchical clustering using complete linkage on a (1 – Pearson *r*) distance matrix was applied to identify patterns of co-regulation among components.

### Methylation-based leukocyte fraction estimation

Blood cell–type proportions were estimated from PacBio HiFi–derived CpG methylation profiles. Methylation calling was performed using pb-CpG-tools (v2.3.2), and per-CpG β-values were computed from the long-read methylation probabilities. Cell-type deconvolution was then carried out using the projectCellType_CP function and the FlowSorted.Blood.EPIC reference dataset (Bioconductor), which performs constrained projection of sample methylation profiles onto reference signatures for major leukocyte populations: CD8 T cells, CD4 T cells, natural killer cells, B cells, monocytes, and neutrophils.

### Cognitive function and functional independence

Unless otherwise specified, all phenotypic measures refer to values obtained at baseline. Global cognitive function was assessed using the Mini-Mental State Examination (MMSE). Additional cognitive tests were administered as described previously in Holstege et al.^28^. Missing cognitive test scores, including MMSE scores, were imputed as described in Beker et al.^7^. Only cognitive tests with more observed than imputed values in the cohort were included in the present analyses. Functional independence was assessed using the Dutch version of the Barthel Index (BI) questionnaire^62^, which evaluates independence in activities of daily living.

Unless otherwise specified, all phenotypic measures refer to values obtained at baseline. Global cognitive function was assessed at baseline using the Mini-Mental State Examination (MMSE), which was used in the present study as a measure of general cognitive status. Missing MMSE scores were imputed as described previously^7^. Functional independence was assessed at baseline using the Dutch version of the Barthel Index (BI) questionnaire^49^, which evaluates independence in activities of daily living.

### Plasma biomarkers

AD-related plasma biomarkers were measured using Simoa-based assays, as described previously in Teunissen et al.^63^. Plasma neurofilament light chain (NfL) was available in 252 centenarians with survival follow-up. Of these, 132 also had DNA methylation, MMSE, Barthel Index, and genetic principal-component data available and were included in the direct survival comparisons involving GrimAge and the median-split NfL analysis.

For correlation analyses with GrimAge acceleration, plasma biomarker measurements were restricted to those obtained within 0.2 years of the blood draw used for DNA methylation profiling, yielding a temporally matched subset of 50 centenarians.

### Hematological markers

Hematological markers were obtained from peripheral blood samples collected at baseline in 133 centenarians and analyzed on a Sysmex XN-9000 hematology analyzer at the Central Research Laboratory of Amsterdam UMC, according to standard laboratory procedures. Routine hematological parameters generated by the analyzer, including absolute blood cell counts and red blood cell indices, were used in the analyses. Leukocyte ratios were calculated from absolute cell counts. The neutrophil-to-lymphocyte ratio (NLR) was defined as the absolute neutrophil count divided by the absolute lymphocyte count. Monocyte-to-lymphocyte, immature granulocyte-to-lymphocyte, eosinophil-to-lymphocyte, and basophil-to-lymphocyte ratios (MLR, IGLR, ELR, and BLR, respectively) were calculated analogously.

DNA methylation-based telomere length (DNAmTL) was estimated using the dnaMethyAge package (https://github.com/yiluyucheng/dnaMethyAge).

### Neuropathological assessments

Neuropathological analyses were performed in 62 brains from centenarians included in this study. The median interval between the blood draw used for DNA methylation profiling and death was 2.45 years. Procedures for brain donation, tissue processing, neuropathological evaluation, and quantification of Aβ8–17 load have been described previously (Rohde et al., 2025).

Additional immunohistochemical assessments quantified Aβ40 (Merck Millipore, #MABN11, 1:1000), Aβ42 (Merck Millipore, #MABN12, 1:5000), and hyperphosphorylated tau using AT8 (Ser202/Thr205; Thermo Scientific, #MN1020, 1:800), pTau217 (Thr217; Invitrogen, #44-744, 1:6400), and GT38 (Abcam, #Ab246808, 1:500). Staining followed the previously described protocol, with modifications to antigen retrieval: formic acid alone was used for Aβ40 and Aβ42, whereas pressure-cooker treatment with citrate buffer alone was used for all hyperphosphorylated tau markers. Amyloid-β and hyperphosphorylated tau loads were quantified in Brodmann area 38 (BA38).

### Medical, sociodemographic, and lifestyle phenotypes

Sensory and mobility assessments were conducted through direct observation by the study team, supplemented with information from participants and/or caregivers when necessary. Functional limitations were rated according to the extent to which vision, hearing, or mobility impairments interfered with daily activities.

Blood pressure was measured at baseline. Hypertension was defined as systolic blood pressure >150 mmHg, use of antihypertensive medication, or a diagnosis documented by a general practitioner. Diabetes was defined as HbA1c >43 mmol/mol, use of oral antidiabetic medication or insulin, or a diagnosis documented by a general practitioner. Dyslipidemia was defined as use of cholesterol-lowering medication or a diagnosis documented by a general practitioner. Medical histories were reviewed, and the number of entries in medical records was summarized by diagnostic category.

Height and weight were primarily self-reported, supplemented where available by measurements obtained during study visits or from medical records. Body mass index (BMI) was calculated from height and weight. Smoking history was assessed as ever versus never smoking. Passive smoke exposure during adulthood was assessed at baseline by asking whether participants had been exposed to smoke daily at home or in the workplace.

Educational attainment was self-reported using the Verhage scale, which classifies education into seven levels ranging from no formal education to completion of a university degree.

### Age acceleration calculation and cohort comparison

Age acceleration was calculated as the residual from a linear regression of each clock’s DNAm age estimate on chronological age across the combined centenarian and AD cohorts. Differences in GrimAge acceleration between cohorts were assessed using a Wilcoxon rank-sum test. We additionally calculated centenarians’ mean deviation from GrimAge estimates predicted by the AD-only regression extrapolated to their chronological ages. The 95% confidence interval was obtained from 10,000 bootstrap iterations, resampling participants separately within each cohort and refitting the AD regression in each iteration to account for uncertainty in the reference line.

### Correlation analysis

Associations between GrimAge acceleration and phenotypic measures were assessed using Pearson correlations for continuous and binary variables and Spearman correlations for ordinal variables. Neuropathological analyses were adjusted for the interval between blood collection and death, with additional adjustment for sex in brain weight analyses. Cognitive test analyses were adjusted for educational attainment, and sociodemographic and lifestyle analyses were adjusted for sex. False discovery rate correction was applied separately within each phenotype domain.

### Survival analysis

Survival time for MMSE and DNA methylation markers was defined from baseline to death or censoring (last confirmed alive). For plasma NfL, survival time was defined from biomarker sampling to death or censoring. Cox proportional hazards models were used to assess associations with survival. Clock comparison models were adjusted for sex and the first five genetic PCs. As a sensitivity analysis independent of the AD calibration cohort, Cox models were also fitted with each unresidualized DNAm clock and chronological age entered as separate standardized terms, with adjustment for sex and the first five genetic principal components. Separately, the primary clock-comparison Cox models were additionally adjusted for methylation-estimated leukocyte fractions. Models including GrimAge, MMSE, plasma NfL, or combinations thereof were adjusted for age, sex, education, and the first five genetic PCs. Hematological survival models were adjusted for age, sex and the first five genetic PCs. Each GrimAge component was analysed in a separate Cox model adjusted for chronological age and the first five genetic PCs. These models were then additionally adjusted for methylation-estimated leukocyte fractions. Continuous predictors were standardized (z-scores), except DNAmPACKYRS in the component survival analyses, which was analysed on its original scale. Hazard ratios represent the effect of a one-standard-deviation increase in standardized predictors and a one-unit increase in DNAmPACKYRS. Kaplan–Meier curves were generated using the survival package in R and stratified at the median or tertiles for visualization.

### RMST analysis

Restricted mean survival time (RMST) was estimated using the survRM2 package in R, with τ = 8.4 years, corresponding to the longest observed follow-up. RMST models were adjusted for sex, education, and the first five genetic PCs. Separate models were fitted for MMSE and GrimAge acceleration. Participants were ranked according to the model-derived linear predictor, and RMST was compared between the lowest- and highest-risk quintiles (20% each). A joint model including both MMSE and GrimAge acceleration was then fitted, and RMSTs were computed for the same lowest- and highest-risk quintiles of the combined score.

### Clustering and regression analysis of hematological markers

Pairwise correlations among hematological markers were computed using Spearman correlation (pairwise complete observations). To identify groups of related markers independent of correlation sign, hierarchical clustering was performed on a distance matrix defined as 1−∣r∣, such that strongly positive and strongly negative correlations were treated as similar. Clustering was performed using average linkage, and the dendrogram was cut into four clusters. For downstream multivariable modeling, the marker most strongly correlated with GrimAge acceleration within each cluster was selected as a representative. Selected markers were standardized (z-scores) and entered into a multivariable linear regression model with GrimAge acceleration as the outcome.

## Author contributions

Data analyses were performed by Y.Z., with support from M.H., A.S., N.T., and H.H. The 100-plus Study was initiated by H.H. The Amsterdam Dementia Cohort was contributed and overseen by E.G.B.V. Neuropsychological assessment and cognitive data collection were performed by M.G., M.v.d.H., and D.D. Simoa plasma biomarker concentration measurements were performed by C.T. and collected by L.L. Neuropathological assessment and data collection were performed by S.K.R. and M.L. Clinical data collection were performed by S.v.d.L. All authors commented on previous versions of the manuscript, and all authors read and approved the final manuscript.

## Acknowledgements

The authors are grateful to all study participants, their family members, the participating medical staff, general practitioners, pharmacists and all laboratory personnel involved in patient diagnosis, blood collection, blood biobanking, DNA preparation and sequencing. Part of the work in this manuscript was carried out on the Cartesius supercomputer, which is embedded in the Dutch national e-infrastructure with the support of SURF Cooperative. This work was supported by a VIDI grant from the Dutch Research Council (NWO 09150172010083), an ASPASIA grant (015.016.059), and a public-private partnership with TU Delft and PacBio, receiving funding from Health∼Holland, Topsector Life Sciences & Health (PPP-allowance), as well as by Alzheimer Nederland (WE.03-2018-07; WE.03-2025-08); the ADORE Foundation; the EU Joint Programme – Neurodegenerative Disease Research (JPND), through ADPriOMICS (JPND2023-1822-087; ZonMw 10510062320005); the Alzheimer’s Association Alzheimer Disease Strategic Fund (ADSF-23-1175650-C); and the BrightFocus Foundation (A2021031S). H.H. and S.v.d.L. are recipients of ABOARD, a public-private partnership receiving funding from ZonMw (73305095007) and Health∼Holland, Topsector Life Sciences & Health (PPP-allowance; LSHM20106). H.H. also received the Alzheimer Research Prize 2020 from the Hans und Ilse Breuer Stiftung and support from the HorstingStuit Foundation (Contribution-2).

Research of Alzheimer center Amsterdam is part of the neurodegeneration research program of Amsterdam Neuroscience. Alzheimer Center Amsterdam is supported by Stichting Alzheimer Nederland and Stichting Steun Alzheimercentrum Amsterdam. The clinical database structure was developed with funding from Stichting Dioraphte.

## Data availability

The data that support the findings of this study are not publicly available due to privacy and ethical restrictions, as they contain information that could compromise participant confidentiality. Data are, however, available from the 100-plus Study upon reasonable request and subject to a data sharing agreement.

The analysis code used to generate the results and figures presented in this study is publicly available on GitHub at https://github.com/holstegelab/clock_paper.

