## Supplementary Information for "Peripheral Epigenetic Aging Predicts Survival Beyond Brain Aging-Related Biomarkers in Centenarians"

**Supplementary Note 1**

To compare DNA methylation values obtained from PacBio long-read sequencing with those from the EPIC array, we analyzed CpG sites shared between the two platforms in one centenarian sample (global coverage = 46.87X). Of the 865,821 CpG sites that can be measured with EPIC array, 845,089 were also present in the PacBio data, showing a high overall correlation (r = 0.96; Supplementary Figure 1A). Among the 20,732 CpG sites missing in the PacBio dataset, 19,881 can be mapped to alternate contigs, 407 had fewer than four mapped reads, and 350 lacked a recognizable CpG dinucleotide in at least one of the sequencing reads, potentially due to germline or somatic variation, or technical artifacts.

To further assess the impact of read depth on methylation estimates, we randomly selected 10,000 sites and sampled 5, 10, 15, and 20 sequencing reads per site. We then compared the methylation values estimated from the sampled reads to the corresponding values from the EPIC array (Supplementary Figure 1). Across all CpG sites, methylation measurements from PacBio and the EPIC array show a strong linear relationship. As read coverage increases, the PacBio estimates converge even more closely with the EPIC array values, indicating higher correlation at higher coverage levels. Most samples had a global coverage of approximately 13 - 20X (median = 15.35; 25th percentile = 12.74; 75th percentile = 20.10), and our read-sampling experiments indicate that, at this depth, the PacBio methylation estimates maintain a high correlation with the EPIC array values in this coverage range.

**Supplementary Note 2**

In this study, we used 246 younger Alzheimer's Disease (AD) patients as the calibration cohort. The AD cohort was used only to extend the chronological-age range for estimating the original age-acceleration residuals; AD participants were not included in any centenarian survival or phenotype association analysis. Notably, while AD is often assumed to be associated with accelerated epigenetic aging, results from peripheral blood studies are mixed. In ADNI, age advancement estimated using the Horvath, Hannum, PhenoAge and GrimAge clocks did not differ significantly between cognitively normal participants, those with MCI and those with dementia[1]. Similarly, longitudinal analyses in Betula and cross-sectional comparisons in AIBL found no significant differences in blood-based Hannum, Horvath or PhenoAge measures between AD cases and cognitively healthy controls[2]. Sibbett et al. initially observed an inverse association between GrimAge acceleration and incident dementia, but this association was attenuated and no longer significant after adjustment for smoking[3] . Conversely, some longitudinal analyses have linked higher blood-based epigenetic age measures to subsequent cognitive decline or progression to MCI or AD[4], and to incident dementia[1]. These findings indicate that associations vary across clocks, cohorts and analytical approaches, and that cross-sectional differences by diagnostic status need not parallel associations with subsequent disease progression. Nonetheless, findings across cohorts remain heterogeneous.

In our calibration cohort, the DNAm GrimAge was highly correlated with chronological age with a Pearson correlation of 0.82 (p = 4.32E-62), indicating that the chronological-age relationship was preserved within the AD cohort. To determine whether the principal survival finding depended on the use of this cohort for residualization, we additionally fitted Cox models restricted to centenarians, including the unresidualized DNAm clock value and chronological age as separate standardized terms. DNAm GrimAge remained strongly associated with mortality (HR = 1.40, 95% CI: 1.21-1.63, adjusted p = 3.55E-05), closely matching the estimate obtained using the original age-acceleration residual (HR = 1.41, 95% CI: 1.22-1.63, adjusted p = 2.14E-05; Supplementary Figure 6A). These results indicate that the association between DNAm GrimAge and mortality was not materially dependent on inclusion of the AD cohort in the residualization model.

**Supplementary Note 3**

To further characterize the biological and clinical correlates of GrimAge acceleration in centenarians, we tested associations with neuropathological measures, cognitive performance, AD-related plasma biomarkers, BMI, education, smoking history, and medical-history variables. In addition, we examined whether individual DNAm GrimAge component scores were associated with these non-survival phenotypes. Full results are provided in Supplementary Tables 2–6 and Supplementary Data 1.

Among neuropathological measures, the strongest association was observed between higher GrimAge acceleration and lower cortical Aβ40 load in Brodmann area 38 (BA38) (Pearson r = -0.47, p = 2.54E-04, adj p = 4.58E-03). Several additional neuropathological measures, including AD neuropathologic change score, atherosclerosis, total Aβ load in BA38, and hyperphosphorylated tau measures detected by AT8, pTau217, and GT38, showed nominally significant negative associations with GrimAge acceleration, but none remained significant after multiple-testing correction.

For cognitive phenotypes, GrimAge acceleration showed nominal positive correlations with digit span forward (Pearson r = 0.15, p = 1.77E-02, adj p = 1.33E-01) and digit span backward (Pearson r = 0.16, 1.07E–02, adj p = 1.33E-01). However, these associations did not survive multiple-testing correction, and GrimAge acceleration was not significantly associated with global cognition or other cognitive test measures.
GrimAge acceleration also did not show significant associations with AD-related plasma biomarkers, BMI, height, weight, smoking history, passive smoke exposure, or Verhage education level after correction for multiple testing. Among medical-history variables, transient ischemic attack/cerebrovascular accident records (Pearson r = -0.18, p = 1.02E-02, adj p = 1.73E-01) and systolic blood pressure (Pearson r = -0.18, p = 4.27E-02, adj p = 3.63E-01) showed only nominal associations. Overall, these results indicate that, outside survival-related and hematological measures, GrimAge acceleration had limited associations with age-related phenotypes in this cohort.

We also evaluated individual DNAm GrimAge component scores in relation to non-survival phenotypes. As described in the main text, GrimAge component scores were generally intercorrelated (Supplementary Figure 5), consistent with the composite nature of the clock. However, component-level associations with non-survival phenotypes were sparse. The clearest association was a positive correlation between DNAmPAI1 and eosinophil count (Pearson r = 0.32, p = 2.02E-04, adj p = 3.72E-02). By contrast, DNAmPACKYRS did not correlate with self-reported smoking history (partial Pearson r adjusted for sex = 0.03, adj p = 9.61E-01) or passive smoking exposure (partial Pearson r adjusted for sex = 0.01, adj p = 9.61E-01). Nevertheless, DNAmPACKYRS was associated with mortality and retained a nominal association after leukocyte adjustment (Supplementary Figure 2A–B). The original GrimAge study also found that this methylation-derived proxy predicted mortality among never-smokers[5] , indicating that its mortality association cannot be interpreted directly as an effect of reported smoking exposure. Survival selection among individuals who reach age 100 may further alter the relationship between smoking exposure and subsequent mortality. Full results for all component-level analyses are provided in Supplementary Data 1.

**Supplementary Figures**

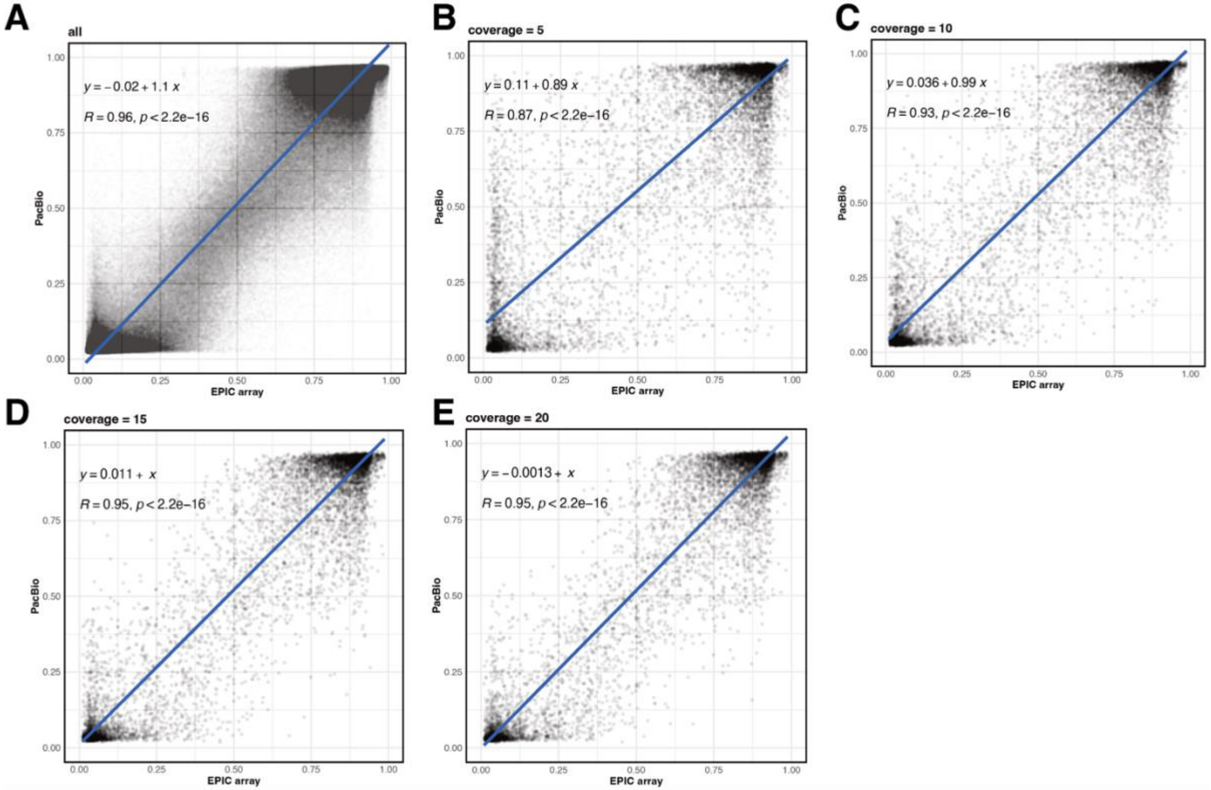


Supplementary Figure 1. (A) Scatter plot comparing DNA methylation values at CpG sites common to the EPIC array and PacBio long-read sequencing in a centenarian sample (global coverage = 46.87X), showing a high concordance between the two platforms (R = 0.96). For 10,000 randomly selected common CpG sites, methylation estimates were calculated by subsampling (B) 5, (C) 10, (D) 15, and (E) 20 reads per site; as the per-site read depth increases, the PacBio-derived methylation values converge more closely with the EPIC array beta values, indicating that a global coverage of 10 - 15X is sufficient for robust analysis.

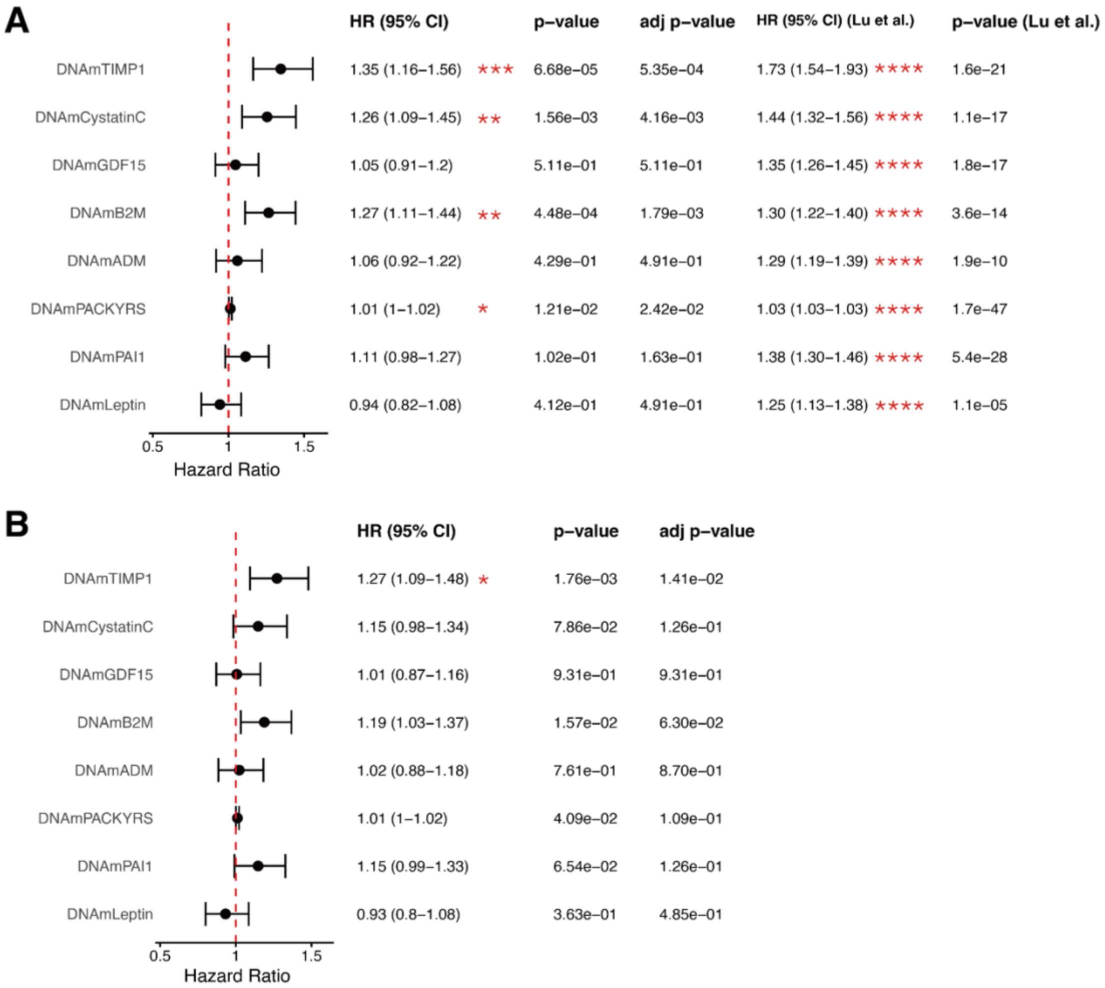


Supplementary Figure 2. Survival associations of DNAm GrimAge components. (A) Cox proportional hazards models for the eight DNAm GrimAge surrogate components, adjusted for chronological age and genetic principal components (n =247, 228 died and 19 censored). All components were standardized except DNAmPACKYRS, which was analyzed on its original scale. Results from the original GrimAge study are shown for comparison[5]. (B) The same models additionally adjusted for methylation-estimated leukocyte composition.

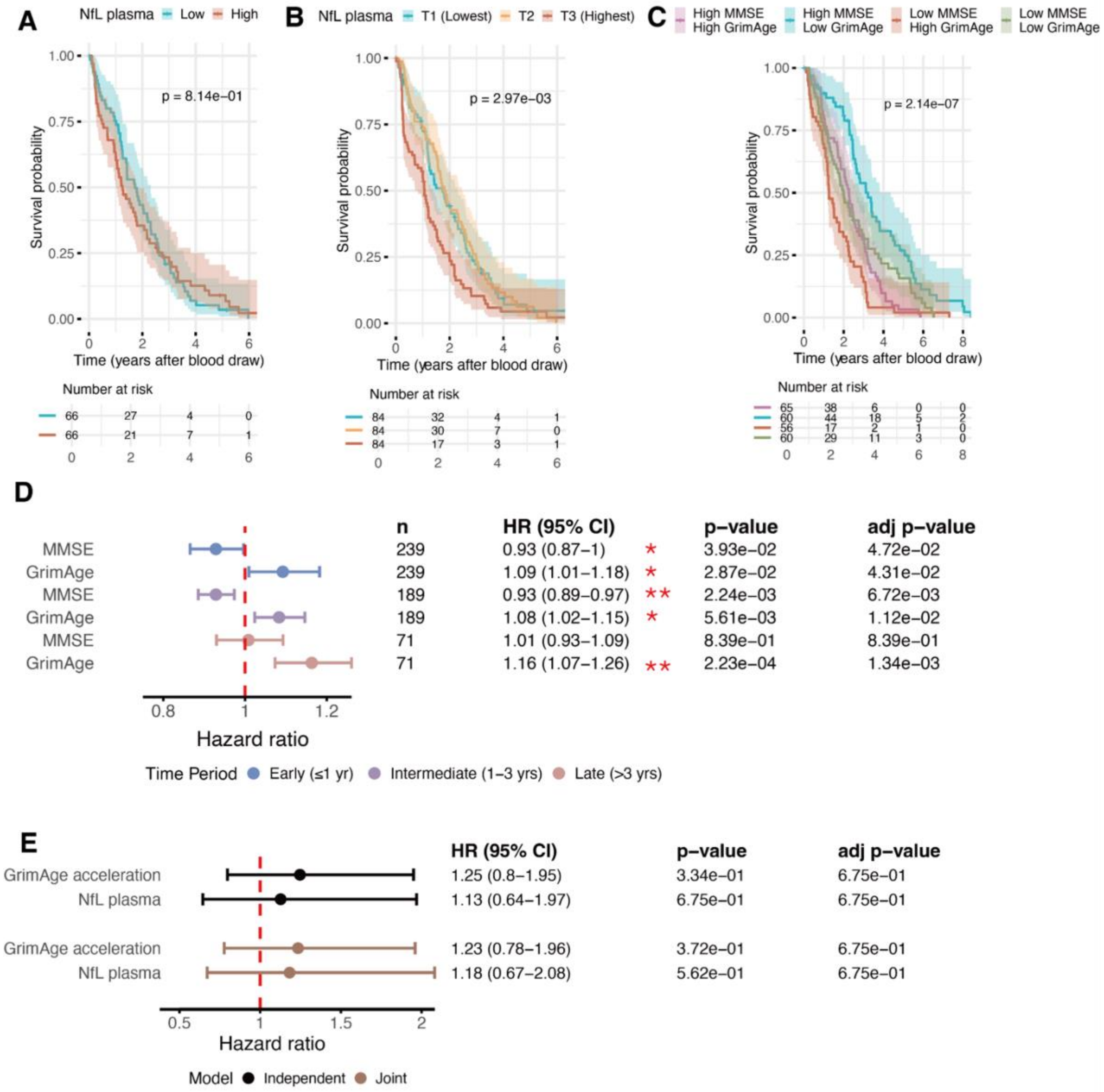
A
NfL plasma Low High
p = 8.14e−01
Survival probability
Time (years after blood draw)
Number at risk
66 27 4 0
66 21 7 1
B
NfL plasma T1 (Lowest) T2 T3 (Highest)
p = 2.97e−03
Survival probability
Time (years after blood draw)
Number at risk
84 32 4 1
84 30 7 0
84 17 3 1
C
High MMSE High GrimAge
High MMSE Low GrimAge
Low MMSE High GrimAge
Low MMSE Low GrimAge
p = 2.14e−07
Survival probability
Time (years after blood draw)
Number at risk
65 38 6 0 0
60 44 18 5 2
56 17 2 1 0
60 29 11 3 0
D
n HR (95% CI) p−value adj p−value
MMSE 239 0.93 (0.87−1) * 3.93e−02 4.72e−02
GrimAge 239 1.09 (1.01−1.18) * 2.87e−02 4.31e−02
MMSE 189 0.93 (0.89−0.97) ** 2.24e−03 6.72e−03
GrimAge 189 1.08 (1.02−1.15) * 5.61e−03 1.12e−02
MMSE 71 1.01 (0.93−1.09) 8.39e−01 8.39e−01
GrimAge 71 1.16 (1.07−1.26) ** 2.23e−04 1.34e−03
Hazard ratio
Time Period Early (≤1 yr) Intermediate (1−3 yrs) Late (>3 yrs)
E
HR (95% CI) p−value adj p−value
GrimAge acceleration 1.25 (0.8−1.95) 3.34e−01 6.75e−01
NfL plasma 1.13 (0.64−1.97) 6.75e−01 6.75e−01
GrimAge acceleration 1.23 (0.78−1.96) 3.72e−01 6.75e−01
NfL plasma 1.18 (0.67−2.08) 5.62e−01 6.75e−01
Hazard ratio
Model Independent Joint

Supplementary Figure 3. Survival analysis with plasma NfL, MMSE, and GrimAge. (A) Kaplan–Meier survival curves based on a median split of plasma NfL in the subset with plasma NfL, GrimAge, MMSE, Barthel Index, and genetic principal-component data (n = 132). (B) Kaplan–Meier survival curves based on plasma NfL tertiles in the larger cohort with plasma NfL, survival follow-up, and genetic principal-component data (n = 252). (C) Kaplan-Meier survival curves were generated based on combinations of median splits of MMSE (25) and GrimAge acceleration (-0.09). (D) Time-stratified Cox regression models examining the associations of MMSE and GrimAge acceleration with mortality during early (0–1 year), intermediate (>1–3 years) and late (>3 years) follow-up. Participants contributed to each interval while alive and under observation. (E) Cox regression models for the associations of GrimAge acceleration and plasma NfL with mortality, restricted to individuals with <0.2 years between the blood draw for plasma NfL measurement and the blood draw for GrimAge estimation (n = 50; 44 deaths, 6 censored). Survival time was defined using the age at blood draw for plasma NfL measurement. Independent model in black, joint model in brown.

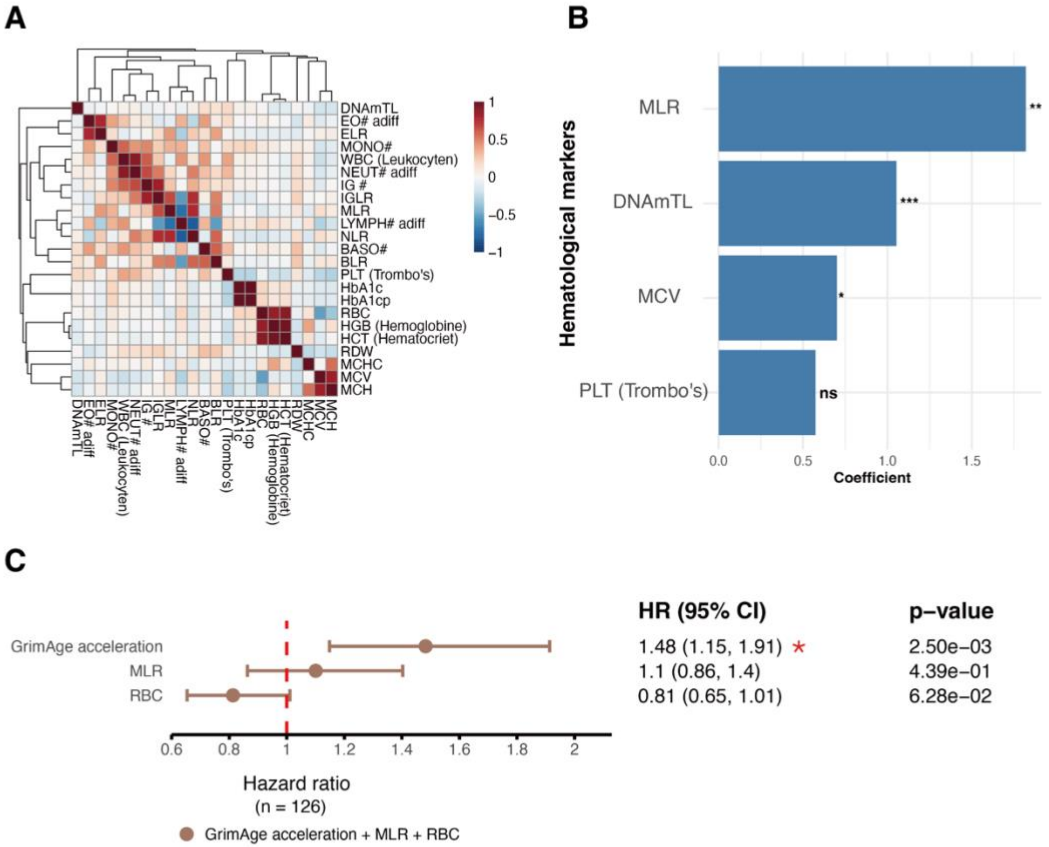


Supplementary Figure 4. Hierarchical clustering of hematological markers and their associations with GrimAge. (A) Spearman correlation analysis followed by hierarchical clustering of hematological markers. Markers were clustered based on their correlation structure, and the representative marker with the highest correlation with GrimAge was selected from each cluster. (B) Linear regression analysis of GrimAge with the four selected representative markers: MLR, DNAmTL, MCV and PLT (Trombo's). Regression coefficients represent the effect size per standard deviation increase in each marker. Significance levels are indicated (*: $p < 0.05$, **: $p < 0.01$, ns = not significant). (C) Cox proportional hazards model including GrimAge acceleration, MLR, and RBC to assess their joint associations with survival (n = 126), adjusted for age, sex and genetic principal components. Hazard ratios (HRs) and 95% confidence intervals are shown.

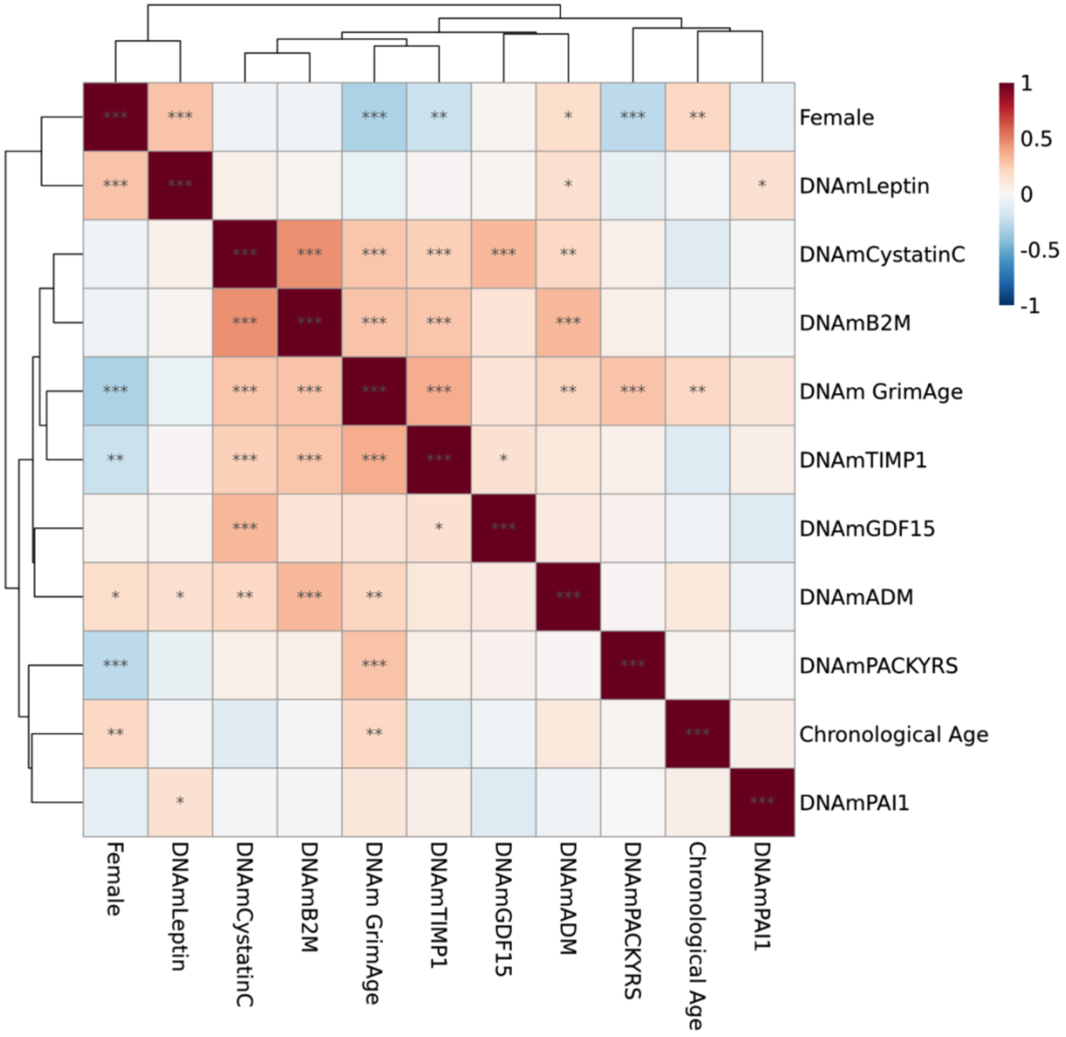


Supplementary Figure 5. Pearson correlation between DNAm surrogates, DNAm GrimAge, chronological age and sex.

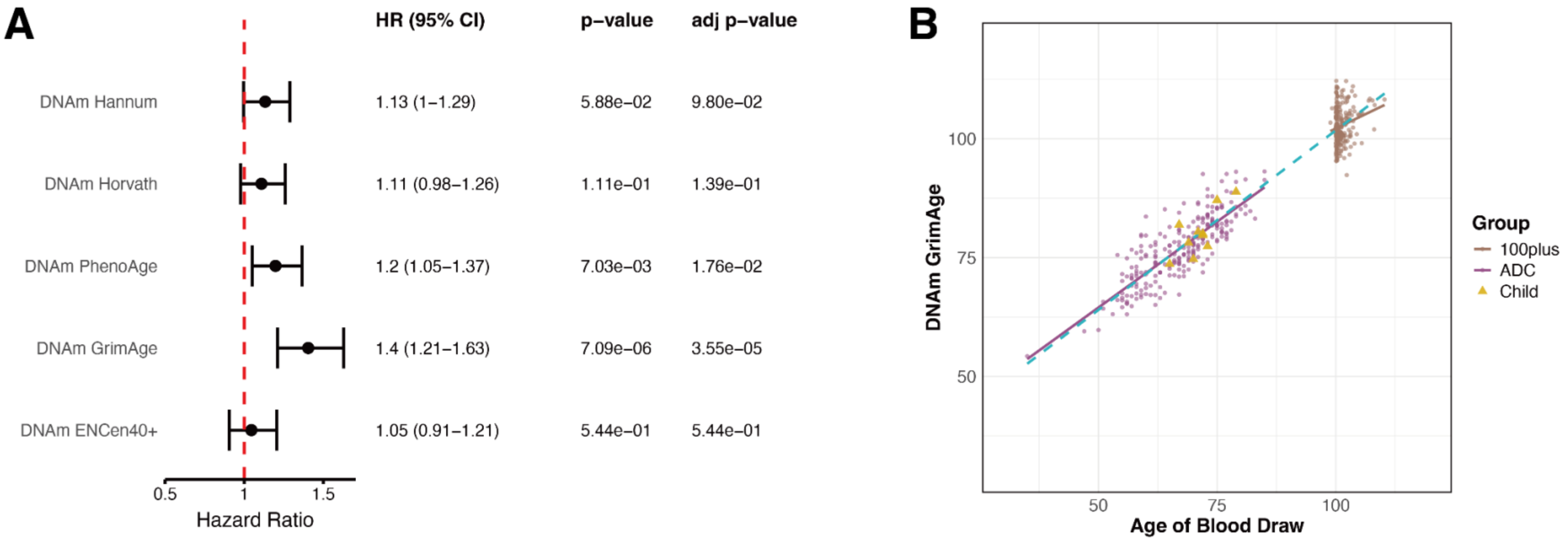


Supplementary Figure 6. (A) Survival associations of unresidualized DNAm clocks after adjustment for chronological age. Cox proportional hazards models were fitted in centenarians with each unresidualized DNAm clock and chronological age included as separate standardized terms. Models were additionally adjusted for sex and the first five genetic principal components. Hazard ratios represent the mortality hazard associated with a one-standard-deviation increase in the respective DNAm clock. Error bars indicate 95% confidence intervals. P-values were corrected across the five clocks using the false discovery rate. (B) Relationship between chronological age at blood draw and DNAm GrimAge in centenarians from the 100-plus Study, participants from the Amsterdam Dementia Cohort, and ten offspring of centenarians. The dashed line represents the linear regression fitted across the combined AD and centenarian samples, which was used to derive the age-acceleration residuals. Offspring are shown as triangles and were not included in the fitted regression.

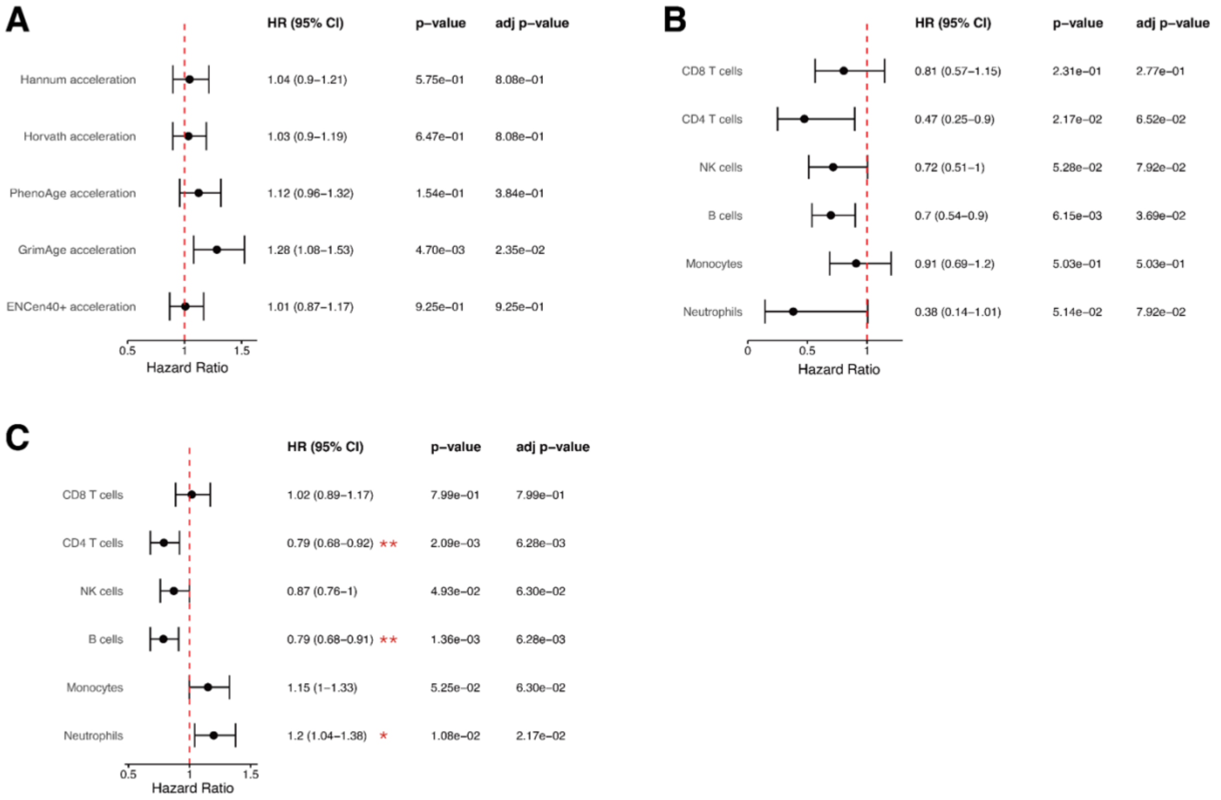


Supplementary Figure 7. (A) Survival associations of epigenetic age acceleration after adjustment for methylation-estimated leukocyte composition. Cox proportional hazards models examining the associations between survival and age acceleration derived from the Horvath, Hannum, PhenoAge, GrimAge and ENCen40+ clocks in centenarians (n = 247, 228 deaths, 19 censored). Models were adjusted for sex, the first five genetic principal components and methylation-estimated major leukocyte populations. All age-acceleration measures were z-score standardized; hazard ratios therefore represent the effect of a 1 SD increase. P-values were FDR-adjusted across the five clocks. (B) Methylation-estimated leukocyte fractions in the GrimAge survival model. Hazard ratios per standard deviation increase in estimated blood-cell fraction, with 95% confidence intervals, for leukocyte fractions included as covariates in the GrimAge–survival Cox model. (C) Survival associations of methylation-estimated leukocyte fractions. Each fraction was analysed in a separate Cox model adjusted for age, sex and the first five genetic PCs.

**Supplementary Tables**

Supplementary Table 1

| Clock | Group | Pearson correlation | p-value | adjusted p-value | MAE | Slope | R squared |
|---|---|---|---|---|---|---|---|
| DNAm Hannum | All | 0.88479745 | 5.96E-165 | 4.47E-164 | 13.247256 | 0.65469902 | 0.78286653 |
| DNAm Horvath | All | 0.86210661 | 4.62E-147 | 1.39E-146 | 18.8555571 | 0.64493073 | 0.74322781 |
| DNAm PhenoAge | All | 0.8780751 | 2.78E-159 | 1.39E-158 | 13.0571035 | 0.78028423 | 0.77101587 |
| DNAm GrimAge | All | 0.961601 | 7.24E-278 | 1.09E-276 | 6.39875835 | 0.75532908 | 0.92467649 |
| DNAm ENCen40+ | All | 0.86572345 | 1.09E-149 | 4.07E-149 | 10.8588022 | 0.7413249 | 0.7494771 |
| DNAm Hannum | 100plus | 0.05327003 | 4.05E-01 | 4.05E-01 | 18.8588035 | 0.24500918 | 0.0028377 |
| DNAm Horvath | 100plus | 0.0621652 | 3.31E-01 | 3.81E-01 | 24.6554321 | 0.3216995 | 0.00386451 |
| DNAm PhenoAge | 100plus | 0.05810442 | 3.63E-01 | 3.89E-01 | 16.7523468 | 0.32345841 | 0.00337612 |
| DNAm GrimAge | 100plus | 0.20405039 | 1.26E-03 | 1.72E-03 | 3.18119866 | 0.48158908 | 0.04163656 |
| DNAm ENCen40+ | 100plus | 0.10069354 | 1.14E-01 | 1.43E-01 | 14.6945381 | 0.53227332 | 0.01013919 |
| DNAm Hannum | ADC | 0.74104141 | 4.26E-44 | 9.13E-44 | 7.61289746 | 0.62028366 | 0.54914238 |
| DNAm Horvath | ADC | 0.70530466 | 2.52E-38 | 4.20E-38 | 13.0321054 | 0.57251389 | 0.49745466 |

| | | | | | | |
|---|---|---|---|---|---|---|
| DNAm PhenoAge ADC | 0.7076341 | 1.13E-38 | 2.11E-38 | 9.34683901 | 0.73139375 | 0.50074602 |
| DNAm GrimAge ADC | 0.82382811 | 4.32E-62 | 1.08E-61 | 9.62939755 | 0.72285608 | 0.67869276 |
| DNAm ENCen40+ ADC | 0.6873016 | 9.81E-36 | 1.47E-35 | 7.0074739 | 0.79645442 | 0.47238349 |

Supplementary Table 2. Correlation between neuropathology markers in the brain with GrimAge acceleration

| variable | n | method | coef (95% CI) | p value | adj p value |
|---|---|---|---|---|---|
| Brain Weight (g) | 57 | pearson | 0.01 (-0.25, 0.27) | 9.25E-01 | 9.25E-01 |
| ADNC (NIA-AA)[6] | 62 | spearman | -0.29 (-0.50, -0.01) | 2.49E-02 | 7.19E-02 |
| Thal Aβ Stage | 62 | spearman | -0.22 (-0.42, 0.03) | 8.46E-02 | 1.46E-01 |
| Braak NFT Stage | 62 | spearman | -0.22 (-0.45, 0.04) | 8.94E-02 | 1.46E-01 |
| CERAD Neuritic Plaque Score | 62 | spearman | -0.21 (-0.42, 0.04) | 9.82E-02 | 1.47E-01 |
| Braak Lewy Body Stage | 62 | spearman | 0.05 (-0.22, 0.29) | 7.08E-01 | 7.49E-01 |
| TDP-43 Pathology Stage[7] | 62 | spearman | -0.24 (-0.47, 0.04) | 6.72E-02 | 1.34E-01 |

| | | | | | |
|---|---|---|---|---|---|
| Thal CAA Stage | 62 | spearman | 0.07 (-0.21, 0.34) | 5.90E-01 | 6.63E-01 |
| Hippocampal Sclerosis | 61 | pearson | -0.11 (-0.36, 0.15) | 3.94E-01 | 4.73E-01 |
| Infarcts | 62 | pearson | -0.24 (-0.46, 0.01) | 6.17E-02 | 1.34E-01 |
| Atherosclerosis | 61 | spearman | -0.32 (-0.50, -0.10) | 1.40E-02 | 6.03E-02 |
| Atrophy | 62 | spearman | -0.17 (-0.40, 0.09) | 2.04E-01 | 2.62E-01 |
| Aβ Load | 57 | pearson | -0.32 (-0.54, -0.07) | 1.52E-02 | 6.03E-02 |
| Aβ40 Load | 57 | pearson | -0.47 (-0.65, -0.24) | 2.54E-04 | 4.58E-03 |
| Aβ42 Load | 57 | pearson | -0.19 (-0.43, 0.08) | 1.57E-01 | 2.18E-01 |
| AT8 Load | 59 | pearson | -0.31 (-0.53, -0.06) | 1.67E-02 | 6.03E-02 |
| pTau217 Load | 60 | pearson | -0.34 (-0.55, -0.09) | 8.05E-03 | 6.03E-02 |
| GT38 Load | 60 | pearson | -0.29 (-0.51, -0.03) | 2.80E-02 | 7.19E-02 |

Supplementary Table 3. Pearson correlation between cognitive tests and GrimAge acceleration

| variable | n | coef (95% CI) | p value | adj p value |
|---|---|---|---|---|
| MMSE score (imputed) | 241 | -0.04 (-0.16, 0.09) | 5.67E-01 | 8.51E-01 |
| MMSE missing indicator | 241 | 0 (-0.12, 0.13) | 9.68E-01 | 9.68E-01 |
| Digit Span Forward (WAIS-III) | 241 | 0.15 (0.03, 0.27) | 1.77E-02 | 1.33E-01 |
| Digit Span Backward (WAIS-III) | 241 | 0.16 (0.04, 0.29) | 1.07E-02 | 1.33E-01 |
| Digit Span Forward — Maximum Span | 241 | 0.12 (-0.01, 0.24) | 7.07E-02 | 2.12E-01 |
| Digit Span Backward — Maximum Span | 241 | 0.12 (-0.01, 0.24) | 6.83E-02 | 2.12E-01 |
| Category Fluency (DAT) | 241 | -0.12 (-0.24, 0.01) | 6.26E-02 | 2.12E-01 |
| Animal Fluency (1 minute) | 241 | -0.08 (-0.2, 0.05) | 2.22E-01 | 5.54E-01 |
| Key Search Test Score | 241 | 0.02 (-0.11, 0.14) | 7.89E-01 | 9.47E-01 |
| Trail Making Test A — Time | 241 | 0.06 (-0.07, 0.19) | 3.51E-01 | 6.58E-01 |
| Trail Making Test A — Errors | 241 | -0.07 (-0.2, 0.06) | 2.76E-01 | 5.92E-01 |
| VAT A — Trials 1 & 2 | 241 | 0 (-0.12, 0.13) | 9.49E-01 | 9.68E-01 |
| VAT Naming — Cue | 241 | -0.05 (-0.17, 0.08) | 4.56E-01 | 7.61E-01 |
| VAT Naming — Goal Stimulus | 241 | -0.02 (-0.15, 0.11) | 7.48E-01 | 9.47E-01 |

| | | | | |
|---|---|---|---|---|
| Clock Drawing Test — Shulman score | 241 | -0.01 (-0.14, 0.11) | 8.21E-01 | 9.47E-01 |

Supplementary Table 4. Pearson correlation between neurodegeneration related plasma biomarkers and GrimAge acceleration

| variable | n | coef (95% CI) | p value | adj p value |
|---|---|---|---|---|
| Aβ40 in plasma | 50 | 0.13 (-0.15, 0.39) | 3.70E-01 | 5.07E-01 |
| Aβ42in plasma | 50 | 0.04 (-0.24, 0.31) | 7.97E-01 | 7.97E-01 |
| Aβ42/Aβ40 ratio (plasma) | 50 | -0.13 (-0.39, 0.16) | 3.80E-01 | 5.07E-01 |
| pTau181 in plasma | 50 | 0.25 (-0.03, 0.49) | 8.13E-02 | 4.88E-01 |
| NfL in plasma | 50 | 0.12 (-0.17, 0.38) | 4.23E-01 | 5.07E-01 |
| GFAP in plasma | 50 | -0.2 (-0.45, 0.09) | 1.71E-01 | 5.07E-01 |

Supplementary Table 5. Correlation between sociodemographic and lifestyle factors and GrimAge acceleration

| variable | n | method | coef (95% CI) | p value | adj p value |
|---|---|---|---|---|---|
| verhage education level | 241 | spearman | 0.09 (-0.04, 0.21) | 1.88E-01 | 5.63E-01 |
| smoker | 238 | pearson | -0.03 (-0.14, 0.1) | 6.76E-01 | 9.50E-01 |
| adult life passive smoking | 223 | pearson | -0.02 (-0.14, 0.11) | 7.92E-01 | 9.50E-01 |
| adult life height | 201 | pearson | 0 (-0.13, 0.13) | 9.63E-01 | 9.63E-01 |

| | | | | | |
|---|---|---|---|---|---|
| adult life weight | 205 | pearson | 0.07 (-0.07, 0.25) | 3.20E-01 | 6.41E-01 |
| adult life BMI | 188 | pearson | 0.13 (-0.03, 0.28) | 7.59E-02 | 4.55E-01 |

Supplementary Table 6. Correlation between other medical related phenotypes and GrimAge acceleration

| variable | n | method | coef (95% CI) | p value | adj p value |
|---|---|---|---|---|---|
| sight | 233 | spearman | 0.04 (-0.09, 0.16) | 5.81E-01 | 7.06E-01 |
| hearing | 236 | spearman | 0.1 (-0.02, 0.24) | 1.17E-01 | 4.45E-01 |
| mobility | 221 | spearman | -0.02 (-0.15, 0.1) | 7.38E-01 | 7.84E-01 |
| TIA / CVA (record) | 212 | pearson | -0.18 (-0.3, -0.04) | 1.02E-02 | 1.73E-01 |
| heart disease (record) | 241 | spearman | 0.09 (-0.04, 0.22) | 1.57E-01 | 4.45E-01 |
| musculoskeletal disease (record) | 241 | spearman | -0.06 (-0.19, 0.07) | 3.68E-01 | 6.26E-01 |
| sensory disease (record) | 241 | spearman | 0.05 (-0.07, 0.18) | 4.58E-01 | 6.54E-01 |
| autoimmune disease (record) | 241 | spearman | -0.08 (-0.21, 0.07) | 2.39E-01 | 5.07E-01 |

| | | | | | |
|---|---|---|---|---|---|
| cardiovascular disease (record) | 241 | spearman | 0.04 (-0.09, 0.17) | 4.97E-01 | 6.54E-01 |
| tumour (record) | 241 | spearman | 0.1 (-0.03, 0.22) | 1.41E-01 | 4.45E-01 |
| other disease (record) | 241 | spearman | 0.03 (-0.1, 0.16) | 6.24E-01 | 7.07E-01 |
| systolic blood pressure | 132 | pearson | -0.18 (-0.34, -0.01) | 4.27E-02 | 3.63E-01 |
| diastolic blood pressure | 132 | pearson | -0.11 (-0.28, 0.06) | 1.97E-01 | 4.78E-01 |
| hypertension | 204 | pearson | -0.05 (-0.18, 0.09) | 5.00E-01 | 6.54E-01 |
| dyslipidemia | 204 | pearson | 0.07 (-0.07, 0.21) | 3.20E-01 | 6.05E-01 |
| diabetes mellitus type 2 | 204 | pearson | 0.12 (-0.02, 0.26) | 8.20E-02 | 4.45E-01 |
| Barthel Index (self-reported) | 247 | pearson | 0 (-0.12, 0.13) | 9.42E-01 | 9.42E-01 |

**Supplementary Data 1.** Associations of individual DNAm GrimAge component scores with hematological markers and other phenotypes in centenarians.

Correlation results for the eight DNAm-based protein and smoking surrogates underlying the original GrimAge clock. Separate worksheets report associations with hematological markers, neuropathological measures, cognitive performance, plasma biomarkers, lifestyle factors, and medical and functional phenotypes. Each worksheet provides the phenotype, DNAm surrogate, sample size, correlation coefficient with its 95% confidence interval, unadjusted P value, and false discovery rate–adjusted P value. Sample sizes vary according to data availability. Statistical procedures and covariate adjustments are described in the Methods.

**Supplementary Data 2.** Numerical results underlying the hematological and GrimAge component analyses.

Numerical results supporting Figure 3A and Supplementary Figures 4A–B and 5. Separate worksheets provide correlations between hematological markers and GrimAge acceleration (Figure 3A); pairwise Spearman correlations among hematological markers and DNAm-estimated telomere length, together with cluster assignments and dendrogram order (Supplementary Figure 4A); results from the joint linear regression of GrimAge acceleration on the selected representative markers (Supplementary Figure 4B); and pairwise correlations among DNAm GrimAge, its surrogate components, chronological age, and sex (Supplementary Figure 5). Results include correlation coefficients, confidence intervals, sample sizes, unadjusted and false discovery rate–adjusted P values, or regression estimates and model statistics, as applicable.
